# A Consensus-Driven Stacking Ensemble Framework for Interpretable Cardiovascular Risk Prediction and Clinical Deployment

**DOI:** 10.64898/2026.05.18.26352989

**Authors:** Shafak Shahriar Sozol, Bipul Chandra Dev Nath, F. M. Shafiullah Fahim, Nusrat Nizam Suzana, Jannatul Ferdous Mirza, Syed Ahmmed, Fatima-Tuz Zohra, Abu Hena Abid Zafr, Mohammed Nasir Uddin, M. Rubaiyat Hossain Mondal, Abu Sayed Md. Latiful Hoque

## Abstract

Machine learning (ML) is being considered to help diagnose cardiovascular diseases (CVD). Still, challenges like inconsistent and limited datasets, limited infrastructure, and global inequalities lead to the need for a reliable and practicable ML solution. This paper presents an ML-driven framework for predicting CVD risk scores and classifying status. Several data preprocessing techniques, including multiple imputation by chained equations (MICE), outlier removal, are considered. In addition, hyperparameter tuning is performed with the GridSearchCV tuning technique. Moreover, a consensus-driven five-feature selection method is applied to identify optimal predictors. The dataset used in this study contains healthcare records related to future CVD risk scores, comprising 1,529 patient records with 22 features. The optimized stacked ensemble model is applied to the dataset and achieves a cross-validated coefficient of determination value of 98.13% for CVD risk score regression. Comparative evaluation with other ML models confirmed improved accuracy, efficiency, and interpretability. The explainable AI technique SHAP is applied to interpret predictions and highlight key risk factors. Moreover, a deployment-ready web platform with multi-role access has been developed that demonstrates clinical applicability. The proposed framework offers a reliable and interpretable tool for early detection of CVD and personalized risk assessment. In the future, this work can be extended to integrate longitudinal data, medical imaging, and deep learning to improve generalizability and strengthen real-world impact.

## 1 Introduction

Cardiovascular disease (CVD) remains a major global health burden. It causes 17.9 million deaths each year and accounts for 32% of global mortality [1]. Hence, early detection and reliable risk stratification are essential to reduce morbidity and mortality. In addition, CVDs also impose socioeconomic costs through medical expenditures, productivity loss, and long-term disability. Therefore, there is a need to develop an effective management system that includes continuous treatment, routine monitoring, lifestyle changes, and coordinated care [2]. However, digital health frameworks and artificial intelligence (AI) provide new opportunities for CVD prevention, diagnosis, and management. In particular, Machine learning (ML), which is a subset of AI, enables data-driven risk predictions supporting more informed clinical decisions [3]. Moreover, hybrid and ensemble ML approaches improve accuracy by combining base models [4]. Additionally, transparency and interpretability are key for clinical adoption. Therefore, explainable AI (XAI) methods strengthen trust in cardiovascular care [1]. However, practical deployment still faces challenges due to inconsistent data standards, incomplete patient records, limited lab integration, and low patient engagement [5]. Furthermore, CVD risk prediction models rely on datasets from developed countries, limiting their applicability in poor and developing countries due to differences in lifestyle and other environmental factors [6]. At the same time, healthcare systems in both poor and developed nations face limited infrastructure and fragmented workflows. Physicians often lack unified patient records, and a few clinical parameters are missing due to limited laboratory resources. As well, laboratory results are disconnected from clinical systems, and patient participation in data management is minimal. As a result, risk prediction tools from developed countries often fail to generalize to low-resource settings [7]. This gap highlights the need for an integrated CVD platform with adaptive inference capability tailored to local clinical and population characteristics.

To address these challenges, this study proposes a reliable and multi-role CVD management framework for developing nations. It also predicts the CVD risk score integrated with adaptive inference capability. The key contributions of this study are as follows:

I. To develop a robust ML framework for continuous risk score prediction using adaptive gradient boosting models to handle missing data during inference.
II. To implement advanced missing data handling through Multiple Imputation by Chained Equations (MICE) and integrate five feature selection techniques for optimal predictor identification.
III. To optimize model performance with systematic hyperparameter tuning using GridSearchCV, ensuring predictive improvements across all models.
IV. To apply explainable AI (XAI) technique to improve model interpretability and identify risk factor prioritization in decision-making.
V. To develop a deployment-ready clinical platform with role-specific dashboards, privacy-preserving patient identification, and Flask API integration for real-world clinical settings.

The rest of the paper is organized as follows. Section 2 reviews related work on CVD risk prediction and AI in healthcare systems. Section 3 discusses the proposed methodology, including dataset description with exploratory analysis, preprocessing, feature selection, model optimization, evaluation metrics, model explainability, and web implementation. Section 4 presents experimental results, covering performance comparisons, optimization and feature selection impact, SHAP visualizations, benchmarking, and platform demonstrations. Section 5 concludes the study.

## 2 Related Works

Recent reports from the American Heart Association confirm that CVD is the leading cause of death in the United States [1]. Therefore, the need for improved early diagnosis and prediction is crucial. ML has appeared as a powerful tool for CVD diagnosis [2, 3]. Typically, CVD prediction is approached in two ways. First, continuous risk score estimation using regression models. The other predicts binary disease presence using classification algorithms. However, recent studies have examined diverse ML techniques for CVD prediction. Alsabhan and Alfadhly [6] evaluated ML and deep learning algorithms for heart disease prediction. They found that ensemble methods generally outperformed base models. In addition, Tuly et al. [8] applied a stacking ensemble combining MLP, XGBoost, Random Forest (RF), KNN, and Logistic Regression (LR) to detect CVD based on lifestyle factors. Olawade et al. [9] showed that Bald Eagle Search Optimisation improved coronary artery disease classification by using feature selection. They explored that the RF model achieved superior performance compared with conventional risk assessments (71–73%). Subsequently, Huang et al. [10] developed a framework using CHARLS? data, in which gradient-boosting models such as LightGBM (AUC = 81.8) and RF (AUC = 81.5) outperformed traditional approaches. A major challenge in real-world applications is incomplete patient data. Most ML models lack adaptive inference capability, meaning they cannot generate reliable predictions when some of the input features are missing [11]. This limitation hinders deployment, as incomplete records are common due to test unavailability, cost constraints, or emergency situations [12]. To address this gap, we require ML models that can handle missing data while maintaining performance. Thereafter, Si et al. [13] predicted postoperative stroke risk in CAD patients using MIMIC IV data. They applied integrated feature selection, reducing 35 features to 14 predictors. CatBoost achieved the best performance (AUC = 84.86), and SHAP identified the Charlson Comorbidity Index and length of stay as critical predictors. In another study, El Sofany et al. [14] evaluated chi-square, ANOVA, and mutual information techniques for feature selection. XGBoost with ANOVA provided good performance, and a mobile application was developed for real-time prediction with SHAP-based interpretability. Additional studies validated ensemble approaches. For instance, Chandrasekhar and Peddakrishna [15] presented a GridSearchCV optimized soft voting ensemble, improving accuracy to 93.44% on the Cleveland dataset. Moreover, Bhatt et al. [16] reported that a Multilayer Perceptron achieved 95% AUC on a large dataset (n = 70,000). For regression-based risk scoring, Sadiq et al. [17] developed models for dyslipidemia-associated CVD risk prediction in pregnant women. RF achieved 95%, though the sample size was small (n = 112). As well, Tasnim et al. [18] proposed a stacking ensemble for cardiovascular risk estimation using Bangladeshi hospital data (n = 1,529). The model achieved 96%, with SHAP identifying BMI, diabetes, and blood pressure as major contributors. However, neither deployment efficiency metrics nor consensus-based feature selection were evaluated.

Despite these advances, several gaps remain. Most studies focus exclusively on either classification or regression, rarely integrating both for comprehensive risk assessment. Limited attention has been given to deployment-oriented efficiency metrics such as inference time, model size, and training speed. Feature selection often relies on single methods rather than consensus-based approaches. Comprehensive SHAP analysis for regression applications is mostly underexplored. Advanced methods for handling missing values are also absent in recent ML based CVD literature. Most critically, prediction models developed using Western datasets may not generalize to South Asian populations due to differences in genetic predisposition, lipid profiles, and disease onset patterns. Studies [7,19] show that Western tools such as the Framingham Risk Score exhibit reduced accuracy in South Asian populations, underscoring the need for locally developed and validated ML framework.

## 3 Materials and Methods

This study aims to develop an automated system for predicting CVD risks. It could benefit physicians and patients. The subsequent sections provide a brief overview of the materials and methods used to achieve this objective. The whole research methodology for CVD disease prediction model is illustrated in Fig. 1.

**Fig. 1.**
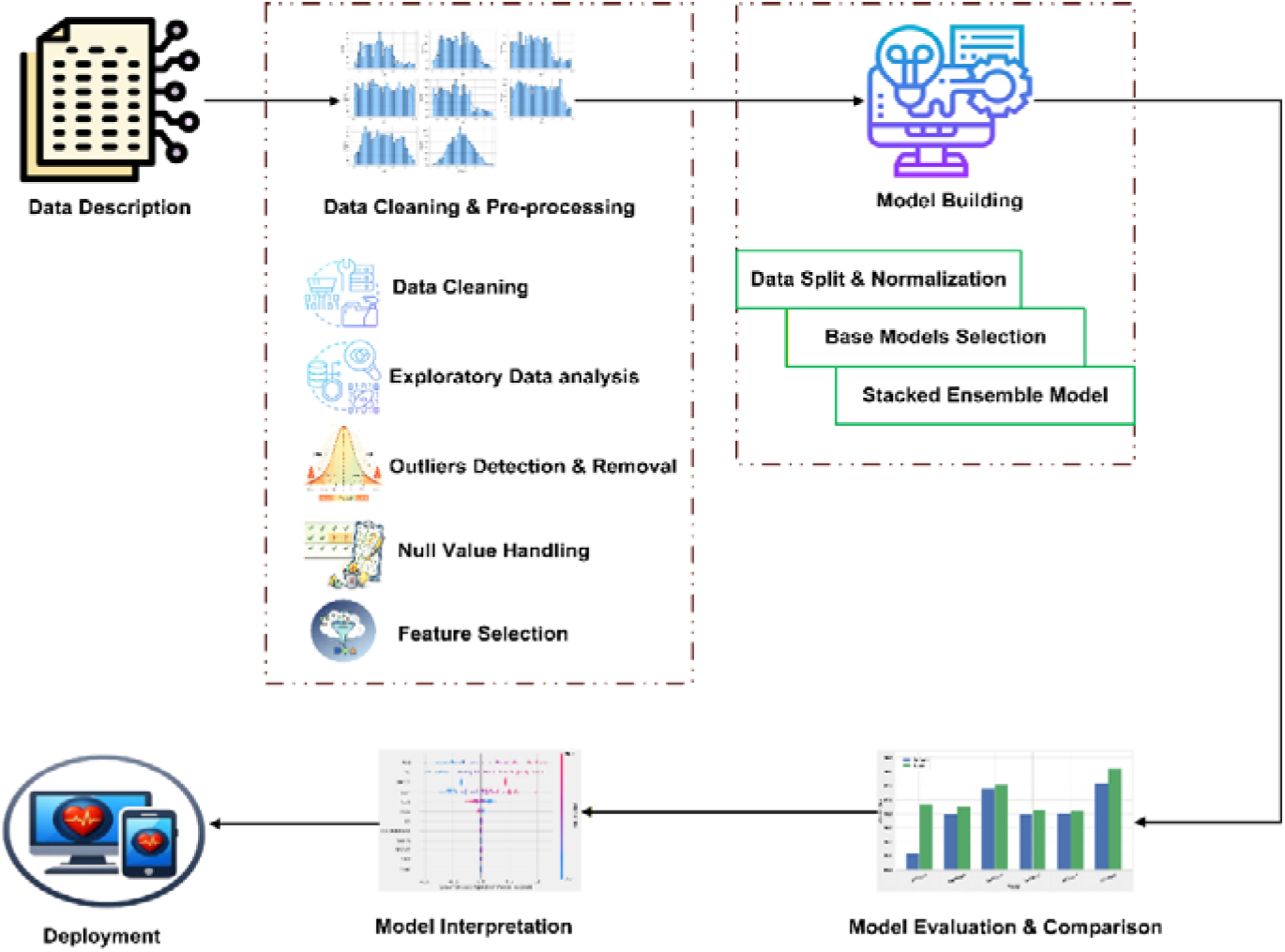
Framework of CVD Detection model

### 3.1 Dataset Overview and Exploratory Analysis

The first dataset considered for this paper was obtained from Jamalpur Medical College Hospital in Jamalpur, Bangladesh, and contains healthcare records related to future CVD risk scores [20]. The dataset comprised 1,529 patient records with 22 features. Features like ‘Height (cm)’, ‘Height (m)’, ‘Weight (kg)’, ‘Blood Pressure (mmHg)’, and ‘CVD Risk Level’ were dropped. These features were either duplicates, less granular representations, or derived. They resulted in redundancy or target leakage. In addition, label encoding technique converted categorical string values into numerical values. Specifically, in this dataset, several categorical variables like Gender (SEX), Blood Pressure Category (BPCAT), Smoking Status (SMK), CVD Risk Score (CVDRISKLVL), Diabetes Status (DMST), Physical Activity (PA), and Family History of CVD (FHxCVD) (variable names are inherited directly from the original dataset nomenclature), contained string based categories. This encoding technique converted categorical variables into integer values. At length, the updated dataset is presented in Table 1.

**Table 1.** Attributes in updated *CVD risk score* dataset and their description.

| Feature Name | Feature Code | Description | Range | Missing Value |
| --- | --- | --- | --- | --- |
| Age | AGE | Age of the individual (Years) | 18–120 | 78 |
| Sex | SEX | Gender (Male/Female) of the individual | — | 0 |
| BMI | BMI | Body Mass Index (kg/m<sup>2</sup>) | 12–60 | 15 |
| Abdominal Circumference | AC | Waist circumference (cm) | 50–150 | 10 |
| Waist-to-Height Ratio | WHtR | Waist-to-height ratio (WHtR) | 0.3–0.8 | 3 |
| Systolic BP | SBP | Systolic Blood Pressure (mmHg) | 70–250 | 0 |
| Diastolic BP | DBP | Diastolic Blood Pressure (mmHg) | 40–130 | 0 |
| Blood Pressure Category | BPCAT | Hypertension Classification | 0-3 | 0 |
| Total Cholesterol | TC | Total cholesterol level (mg/dL) | 100–400 | 73 |
| Fasting Blood Sugar | FBS | Fasting blood glucose level (mg/dL) | 40–300 | 67 |
| HDL | HDL | HDL cholesterol level (mg/dL) | 20–100 | 80 |
| Estimated LDL | LDL | LDL cholesterol level (mg/dL) | 30–300 | 69 |
| Smoking Status | SMK | Current smoking status | 1/0 | 0 |
| Physical Activity | PA | Physical activity level (Low/Moderate/High) | 0-2 | 0 |
| Diabetes Status | DMST | Presence or absence of diabetes mellitus | 1/0 | 0 |
| Family History of CVD | FHxCVD | Family history of cardiovascular disease | 1/0 | 0 |
| CVD Risk Score | CVDRISK | Cardiovascular disease risk score | 0–100 | 69 |

**Note:** 1 = presence, 0 = absence for categorical variables

Distribution plots represent demographic and clinical characteristics (Supplementary Figs. 2a–g). BMI is moderately right skewed, with most participants in the normal to overweight range. In contrast, DBP follows a symmetric pattern, while SBP shows greater variability. Similarly, TC and FBS show mild dispersion. It indicates potential cardiovascular risk in a subset. HDL appears balanced; however, LDL shows wider spread which presents lipid variability. Categorical variables (Supplementary Figs. 2h–l) are evenly distributed across SEX, BMI, WHtR, SMK, and FHxCVD. As a consequence, this supports unbiased group comparisons. In addition, correlation heatmap (Supplementary Fig. 3) shows BMI is strongly associated with CVDRISK (r = 0.62) and SBP indicates a moderate correlation (r = 0.48). Therefore, both features come out as key contributors to CVDRISK.

### 3.2 Outlier Detection and Standardization

Outlier detection is essential in preprocessing as it removes values that deviate sharply from normal distributions. Thereby, it improves model reliability. In this study, the Interquartile Range (IQR) method was applied to numerical variables and filtered records beyond defined bounds [24]. IQR Eq. (1) is as follows:

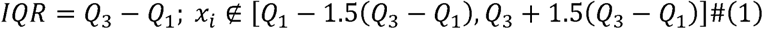

where *Q_1_* and *Q_3_* are the first (25th percentile) and third (75th percentile) quartiles. After filtering, total number of records was reduced from 1,529 to 1,522 samples. Subsequently, standardization technique was applied to numerical features. By following this technique, *AGE, BMI, AC, WHtR, SBP, DBP, TC, FBS, HDL, LDL, and CVDRISK* features were scaled. Thereafter, each feature was transformed to a mean of zero and a standard deviation of one. This scaling ensured all variables were on the same scale to improve comparability.

### 3.3 Missing Value Handling

MICE (Multiple Imputation by Chained Equations) [25] is a method for handling missing values in numerical datasets. Specifically, it predicts missing values iteratively from other features. In this study, imputation was applied only to numerical columns. In addition, *IterativeImputer* from scikit-learn was used with 50 iterations to ensure stable estimates. Afterwards, imputed values were converted to absolute values. Moreover, integer columns were rounded and cast back to primary data type, while float columns were preserved. Finally, the imputed version was used for subsequent analysis.

### 3.4 Feature Selection Technique

A systematic evaluation of multiple feature selection (FS) techniques [26] was conducted to improve the predictive strength of the CVD model. In particular, five FS methods were applied: Pearson correlation, Recursive Feature Elimination (RFE), ANOVA F-test, Elastic Net regularization, and XGBoost feature importance. Pearson correlation FS method identified features having strong linear associations with the target feature. As well, RFE iteratively removed less informative predictors. ANOVA F-test statistically assessed feature relevance. Subsequently, Elastic Net balanced sparsity and stability by using L1-L2 penalty combination, while XGBoost provided feature importance rankings. At length, the integration of filter (Pearson Correlation & ANOVA F-test), wrapper (RFE), and embedded (Elastic Net & XGBoost Importance) FS approaches resulted in a robust, reliable and discriminative framework for CVD risk prediction.

### 3.5 Hyperparameter Optimization

Hyperparameters are model training parameters defined before the execution of ML algorithms [27]. They regulate model behavior and performance; however, improper tuning may yield suboptimal outcomes. In this study, hyperparameter tuning was performed using GridSearchCV technique. Specifically, this technique evaluated every possible parameter combination with 10-fold CV. Thus, it ensured model assessment under diverse configurations. Consequently, the process identified the parameter set that maximized predictive performance.

### 3.6 ML Models

In this study, ML models integrated with adaptive inference capability were selected for CVD risk score prediction. In effect, this inference property enabled prediction during inference even when some input features were missing. However, we used ML algorithms such as Extreme Gradient Boosting (XGBoost) [28], Light Gradient Boosting Machine (LightGBM) [29], CatBoost [30], Histogram based Gradient Boosting (HistGrad) [31], Natural Gradient Boosting (NGBoost) [32], and a Stacking Ensemble [33]. Primarily, the stacking ensemble model combined three tuned models i.e., XGBoost, CatBoost, and LightGBM. These Tuned models were trained separately on the same dataset. Subsequently, their predictions were aggregated through a tuned Ridge Regression model, which learned to optimize outputs for the final prediction.

### 3.7 Model Evaluation Metrics

To ensure fair comparison between ML models, both predictive performance and computational efficiency metrics were evaluated. In this case, error-based metrics like Mean Absolute Error (MAE), Mean Squared Error (MSE), and Root Mean Squared Error (RMSE), along with R-squared (R^2^), were used for CVD risk score prediction. However, lower MAE, MSE, and RMSE metrics indicate smaller prediction errors; conversely, higher R^2^ values show better model performance. Moreover, computational metrics such as training time, inference time, and model size were recorded to assess efficiency and deployment feasibility. As well, 10-fold CV technique was applied to reduce overfitting bias and ensure statistical reliability. Table 2 presents the performance metrics used for model evaluation, covering detection and computational measures.

**Table 2.**
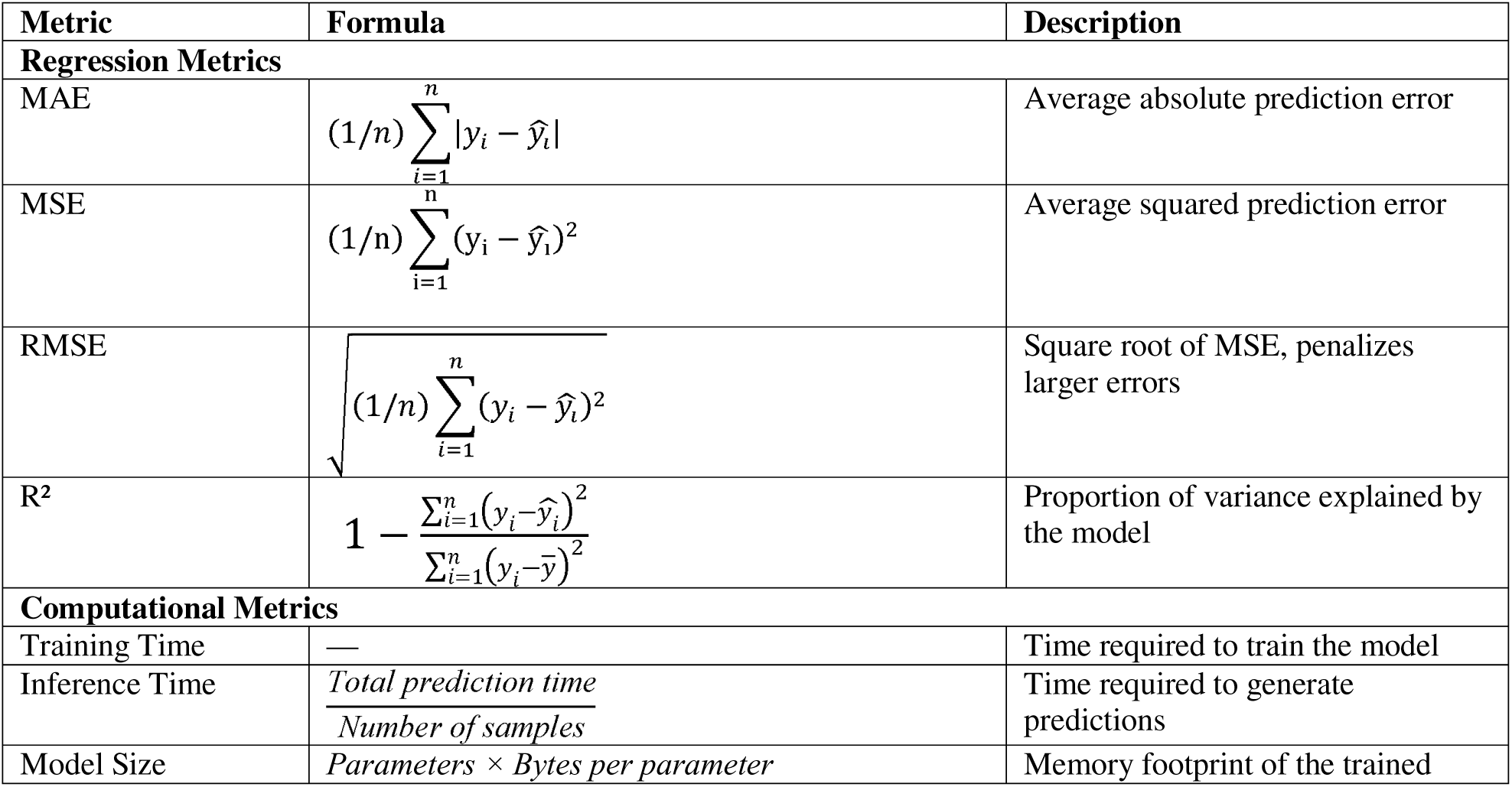
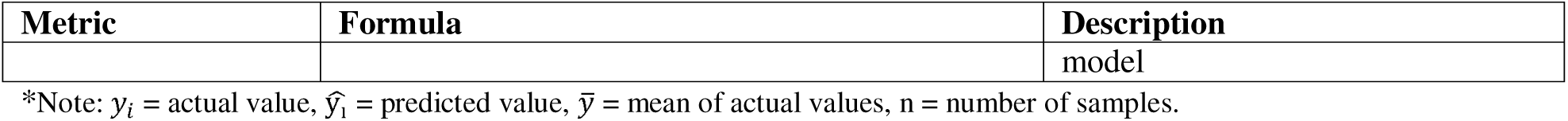
Performance metrics for model evaluation.

### 3.8 SHAP as XAI

To improve model transparency, we applied SHAP (SHapley Additive exPlanations) [34] that was a game theoretic approach for ML models’ feature attribution. In particular, SHAP quantifies each feature contribution to individual predictions. The Shapley value (*ϕ_i_*) for feature *i* is shown in Eq. (2):

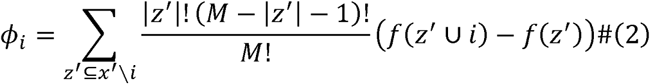

Here, M is the total number of features, z′ represents a subset of features, and f(z′) is the model prediction based on that subset. In this study, *KernelSHAP* was used with 100 training samples as background and SHAP values were computed for 50 test samples. Indeed, feature importance was visualized using global summary plots.

### 3.9 Implementation of Web Application

This section describes the development of a web application for CVD risk prediction. Primarily, web pages of this application were developed using the Laravel MVC architecture. The application can take user input, and the input form contains patients’ demographic details and clinical information, as illustrated in Fig. 2(a) and Fig. 2(b). Furthermore, to ensure faster record retrieval and maintain patient anonymity, a unique patient identification key (PIK) was generated from their demographic information [35].

**Fig. 2.**
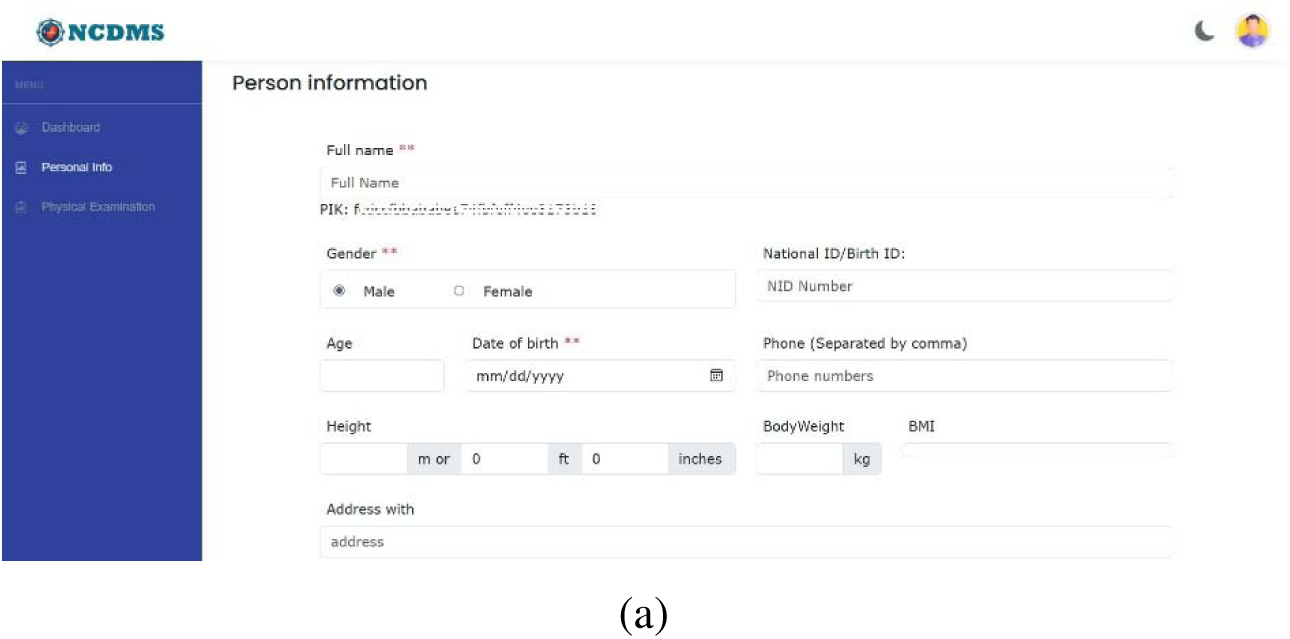
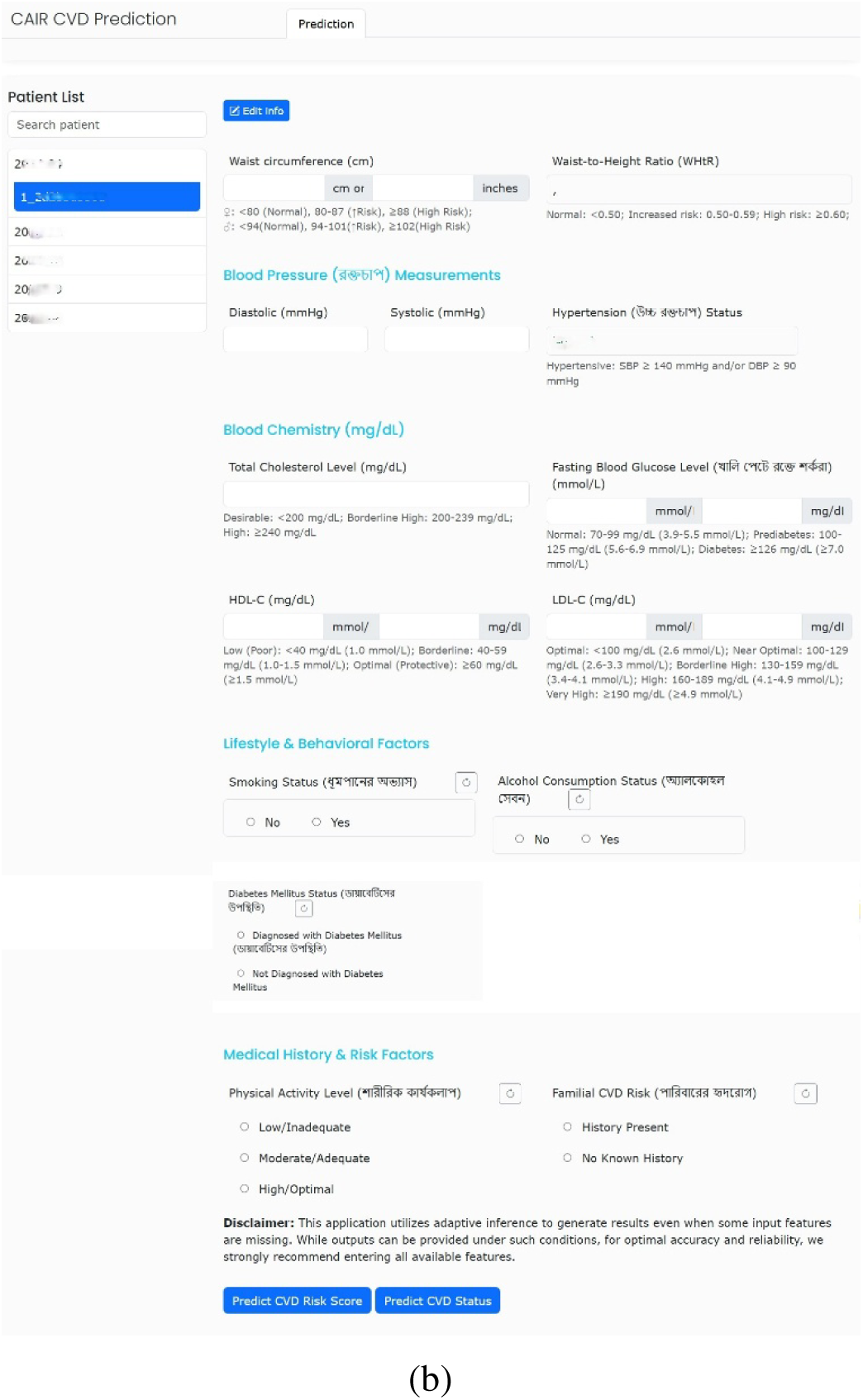
Input user form: (a) Patient demographic, (b) Clinical information

At first, a pickle file of trained model was generated in Kaggle notebook [36]. A dedicated API was then created using the Flask API framework. This Flask API serves as a bridge between the webpage and the ML model. However, when a user inputs values and clicks the predict button, the parameters are sent to the Flask API. Then, the API forwards the inputs to the ML model for CVD risk score prediction. After generating the disease risk score, the results are returned to the webpage via the API. Thus, it allows users to view the output risk score. Therefore, the system uses a unified AI service layer in which the Flask service encapsulates trained predictive model. After that, Laravel validates client input and coordinates with the AI service through RESTful API calls. Subsequently, the ML model returns predictions in a structured format that includes the predicted risk score and its categories. Finally, these outputs are then interpreted and displayed in a clinically interpretable manner. The key architecture of the proposed software is shown in Fig. 3.

**Fig. 3.**
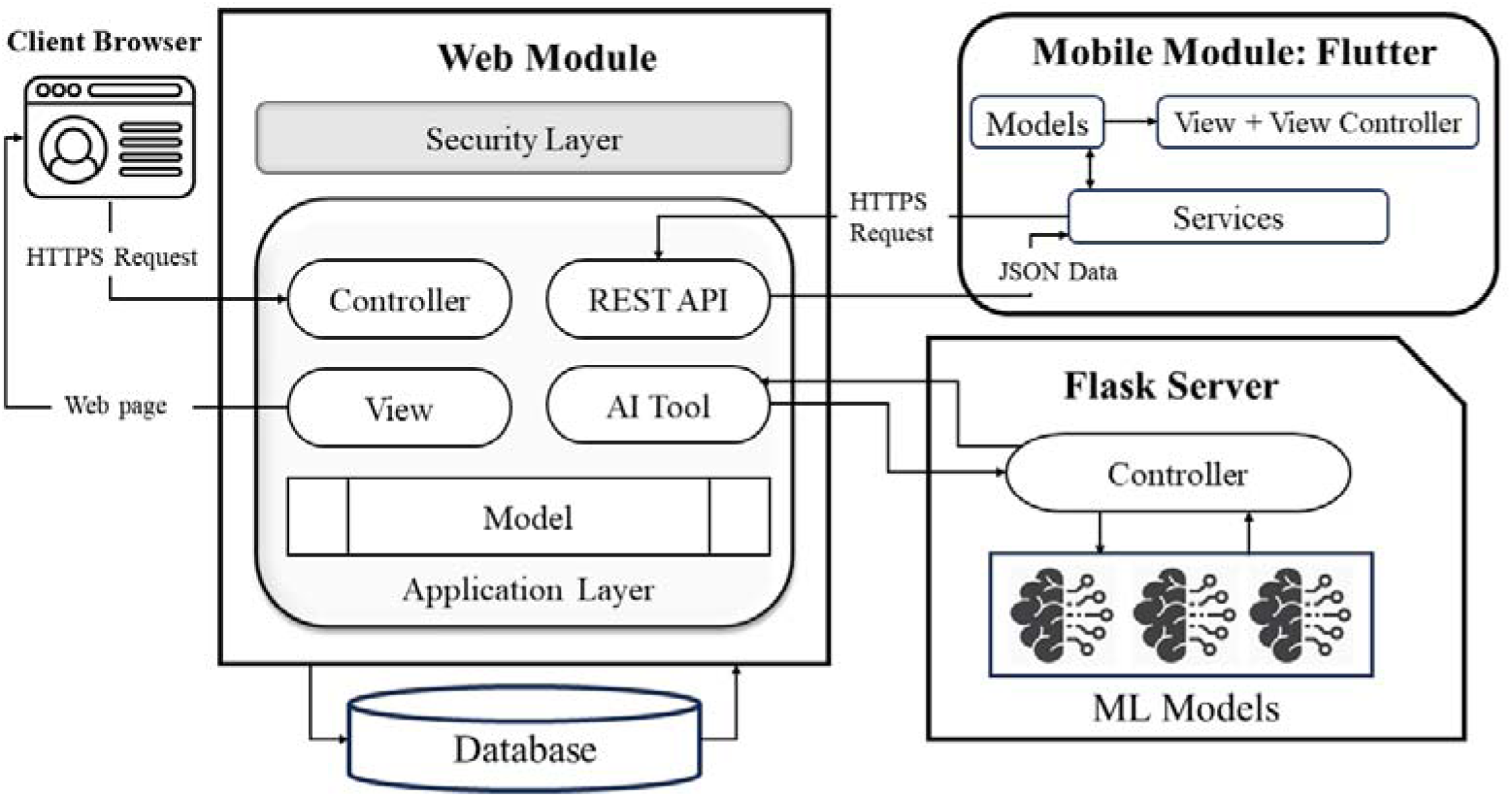
Software architecture

## 4 Experimental Results and Discussions

The performance was assessed across six state-of-the-art ML model regressors with adaptive inference capabilities. Among these model configurations, the best-performing model was identified through robust evaluation metrics. Primarily, the dataset was split into 70% for training and 30% for testing sets. Importantly, this section outlines the experimental setup, comparative performance of each individual model, and final optimized results achieved through stacking generalization.

### 4.1 Environment Setup

In this study, all computational experiments were conducted in a high-performance Kaggle platform. In addition, the system was equipped with an Intel® Xeon® CPU running at 2.00 GHz with four virtual cores, 16 GB of RAM, and a 38.5 MiB L3 cache. The system CPU architecture supported 64-bit processing (x86_64).

### 4.2 Feature Importance Analysis

Based on consensus analysis across five feature selection methods, the primary feature list was refined by evaluating each attribute contribution. Thereafter, this consolidated analysis is presented in Table 3. The initial dataset contained 17 variables, including SEX, AGE, and FHxCVD, but only the most relevant predictors were retained. However, BMI, DMST, SBP, and TC predictors achieved the highest consensus, as they were selected by all five methods. It indicates these features have strong and stable associations with the target outcome. As well, BPCAT and LDL were selected by four methods. DBP and WHtR were retained by three methods and HDL, PA, and SMK were selected by two techniques. In contrast, AC and FBS received minimal support from only one method. Consequently, due to limited consensus, lower ranked features (AC and FBS) were excluded from the final modelling phase. At length, this consensus driven refinement ensures that the final model is constructed using the most robust and consistently validated predictors.

**Table 3.**
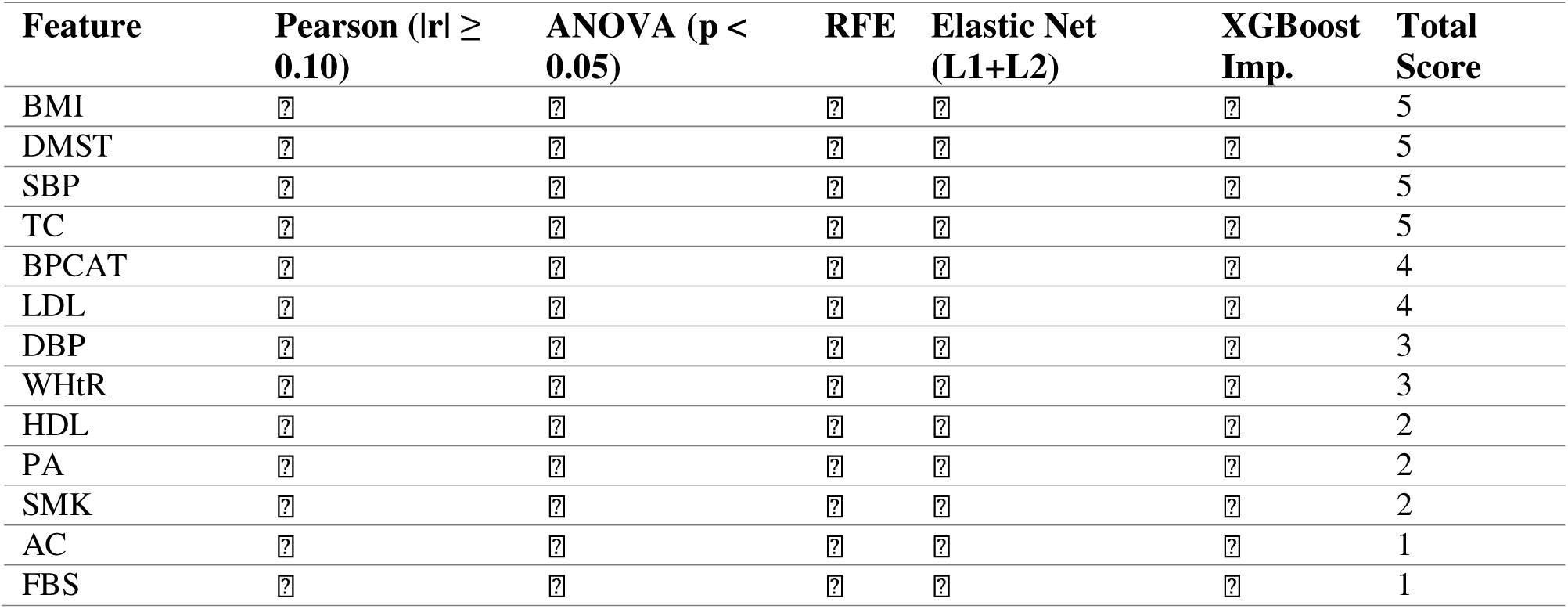
Feature selection consensus.

### 4.3 Hyperparameter Optimization

To improve model predictive performance and optimization, hyperparameter tuning was performed using GridSearchCV with 10-fold CV. The optimal hyperparameter settings for each model are shown in Table 4. XGBoost and LightGBM converged to identical hyperparameter configurations. This identical similarity reflects comparable optimization behavior. In addition, CatBoost and NGBoost preferred lower learning rates of 0.05. CatBoost selected depth 4, whereas NGBoost used 300 estimators with natural gradient descent. HistGrad achieved optimal performance with a maximum depth of 5 and L2 regularization of 0.1. Subsequently, only the meta-learner (ridge) in the stacking ensemble model was tuned. The tuned parameters consistently improved CV accuracy, which supports the validity of the tuning approach.

**Table 4.** Optimal Hyperparameters from GridSearchCV.

| Base Model | Optimal Hyperparameters |
| --- | --- |
| XGBoost | {'learning_rate': 0.1, 'max_depth': 3, 'n_estimators': 200, 'subsample': 0.8} |
| LightGBM | {'learning_rate': 0.1, 'max_depth': 3, 'n_estimators': 200, 'subsample': 0.8} |
| CatBoost | {'depth': 4, 'iterations': 200, 'l2_leaf_reg': 1, 'learning_rate': 0.05} |
| HistGrad | {'l2_regularization': 0.1, 'learning_rate': 0.1, 'max_depth': 5, 'max_iter': 100, 'max_leaf_nodes': 31} |
| NGBoost | {'col_sample': 1.0, 'learning_rate': 0.05, 'minibatch_frac': 1.0, 'n_estimators': 300, 'natural_gradient': True} |
| Stacking (Ridge) | {'final_estimator': Ridge (alpha=0.01, random_state=42), 'final_estimator__alpha': 0.01, 'passthrough': True} |

### 4.3 Results Before FS

The performance of each regression model was evaluated before FS and after optimization. This is shown in Table 5. Therefore, tuned Stacking Ensemble model achieved the highest CV R^2^ of 98.05%, followed by tuned CatBoost and default Stacking Ensemble. Moreover, optimization improved the predictive performance, like Stacking Ensemble from 97.7% to 98.05% and CatBoost from 97.25% to 97.5%. In addition, LightGBM showed the greatest reduction in training time, thereby decreasing from 0.1664s to 0.0518s. CatBoost achieved the fastest inference time of 2.0 ms (for 100 samples) and the smallest model size of 0.07 MB after tuning. However, not all models showed uniform improvement. In particular, XGBoost increased from 95.1% to 96.85%, HistGrad improved from 96.4% to 96.65%, and NGBoost showed only marginal gains despite efficiency improvements. Nevertheless, optimization improved performance overall by reducing error metrics and increasing cross-validated R^2^ scores, as illustrated in Fig. 4.

**Fig. 4.**
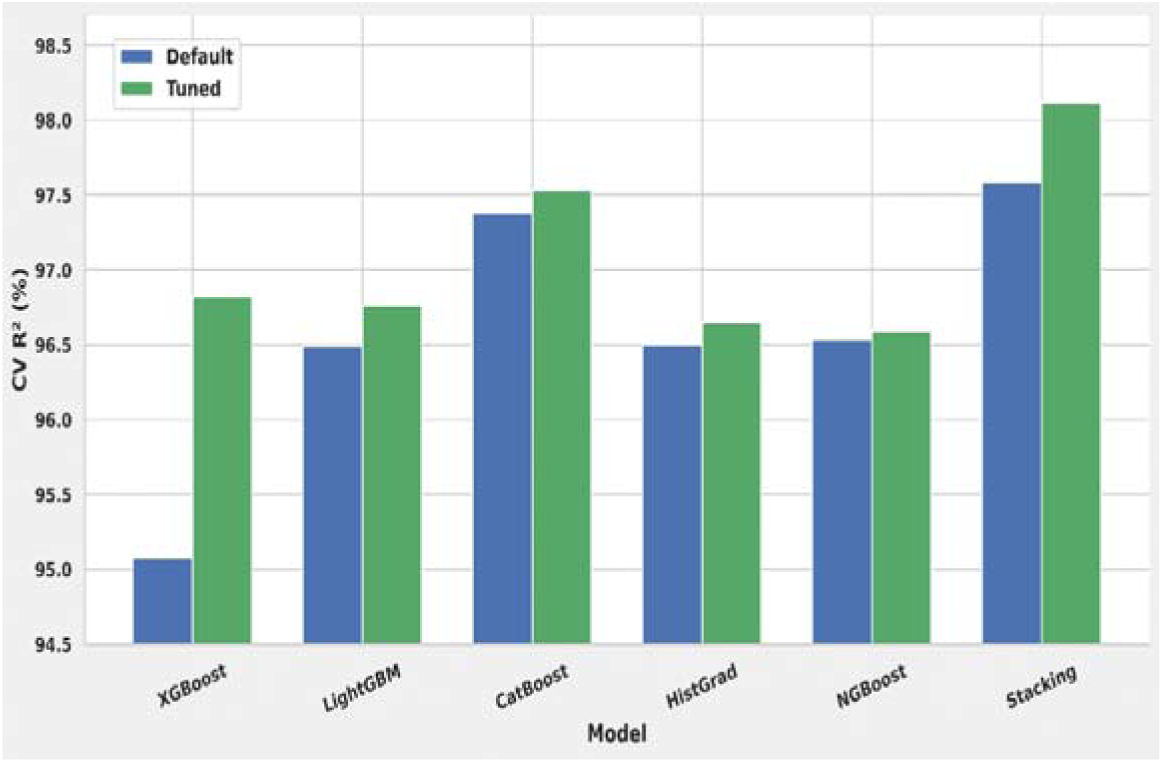
Performance impact of optimization based on CV R^2^

**Table 5.** Default vs Tuned model performance before FS.

| Model | Config. | MAE | MSE | RMSE | $R^2$ (%) | Training Time (s) | Inference Time (ms) | Model Size (MB) |
| --- | --- | --- | --- | --- | --- | --- | --- | --- |
| XGBoost | Default | 0.1486 | 0.0432 | 0.2078 | 95.08 | 0.2156 | 7.6 | 0.3515 |
|  | Tuned | 0.1073 | 0.0276 | 0.1661 | 96.82 | 0.2175 | 5.4 | 0.2251 |
| LightGBM | Default | 0.1213 | 0.0318 | 0.1783 | 96.49 | 0.1664 | 4.8 | 0.2797 |
|  | Tuned | 0.1089 | 0.0268 | 0.1638 | 96.76 | **0.0518** | 3.9 | 0.1466 |
| CatBoost | Default | 0.0860 | 0.0243 | 0.1559 | 97.38 | 1.9808 | 4.7 | 1.0680 |
|  | Tuned | 0.0815 | 0.0222 | 0.1489 | 97.53 | 0.2371 | **2.0** | **0.0705** |
| HistGrad | Default | 0.1255 | 0.0340 | 0.1845 | 96.50 | 0.4963 | 7.7 | 0.3321 |
|  | Tuned | 0.1210 | 0.0327 | 0.1808 | 96.65 | 0.2299 | 5.2 | 0.1906 |
| NGBoost | Default | 0.1167 | 0.0299 | 0.1729 | 96.53 | 7.0244 | 133.0 | 1.3343 |
|  | Tuned | 0.1093 | 0.0272 | 0.1651 | 96.59 | 4.0905 | 83.4 | 0.7830 |
| Stacking | Default | 0.0749 | 0.0210 | 0.1450 | 97.59 | 3.1235 | 16.5 | 0.8853 |
|  | Tuned | **0.0540** | **0.0178** | **0.1335** | **98.12** | 3.5934 | 12.9 | 0.4449 |

Note: Best values in each column are highlighted in bold.

The outcomes from regression models confirm that GridSearchCV effectively identifies optimal hyperparameter settings.

### 4.3 Results After FS

After feature selection, the eleven retained predictors, including BMI, DMST, SBP, TC, BPCAT, LDL, DBP, WHtR, HDL, PA, and SMK were used to evaluate all models under default and tuned configurations. It is shown in Table 6. The tuned Stacking Ensemble achieved the best performance, with a CV R^2^ of 98.13% and the lowest error metrics (MAE: 0.0538, MSE: 0.0178, RMSE: 0.1335). In particular, CatBoost was the most deployment friendly model, which achieved a CV R^2^ of 97.58%, the fastest inference time of 1.2 ms, and a smaller model size of 0.07 MB. Subsequently, LightGBM showed the largest efficiency gain as it reduced training time from 0.1105s to 0.0348s while maintaining competitive accuracy. However, models’ performance improvements varied. XGBoost model increased CV R^2^ from 95.40% to 96.93%, HistGrad rose from 96.66% to 97.78%, while NGBoost delivered only marginal predictive gains despite substantial reductions in training time and model size. Therefore, hyperparameter tuning improved performance, with the tuned Stacking Ensemble model considered as the most reliable configuration following FS, as illustrated in Fig. 5. These results confirm that FE combined with hyperparameter optimization effectively improves model performance and efficiency for CVD risk score prediction.

**Fig. 5.**
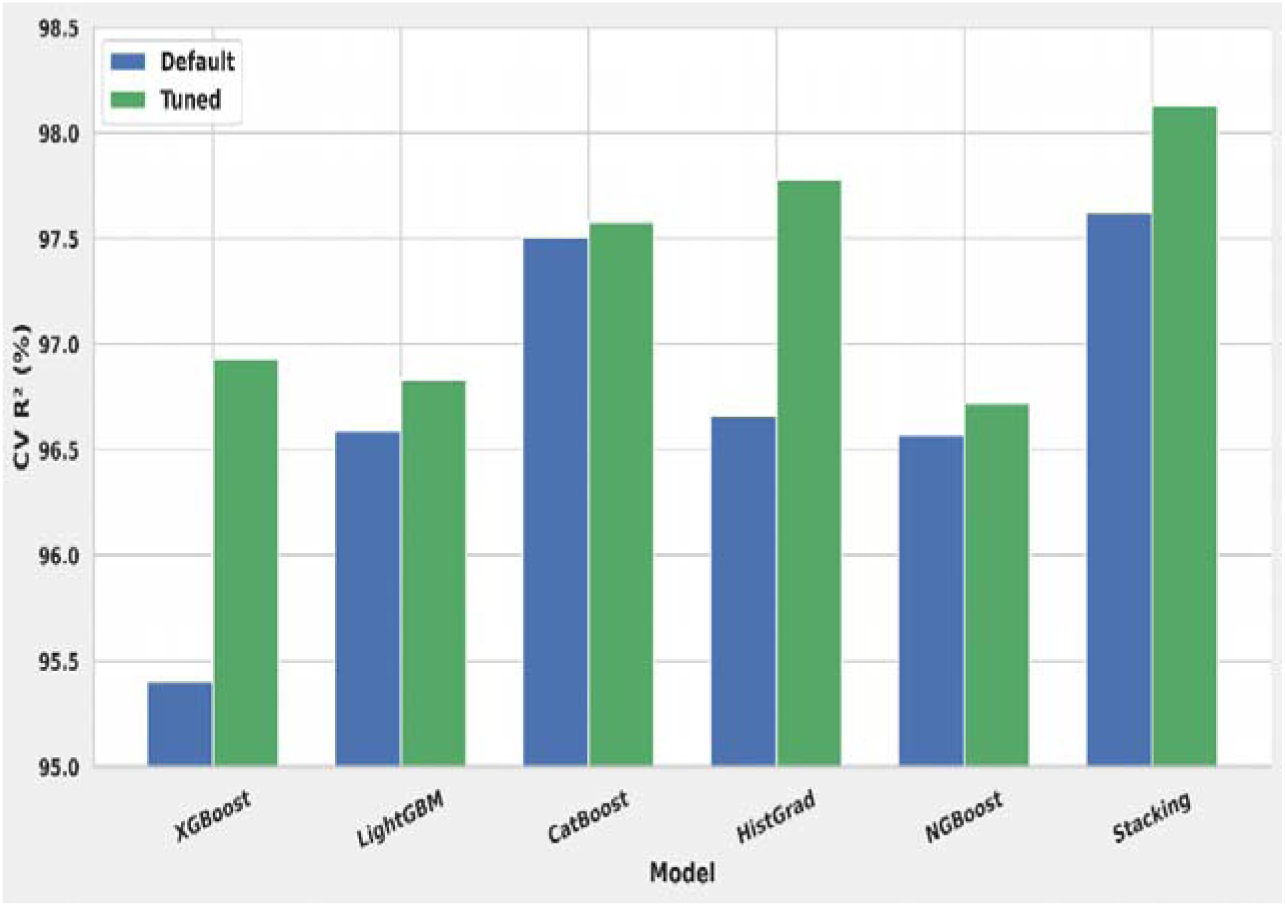
Performance impact of tuning after FE

**Table 6.**
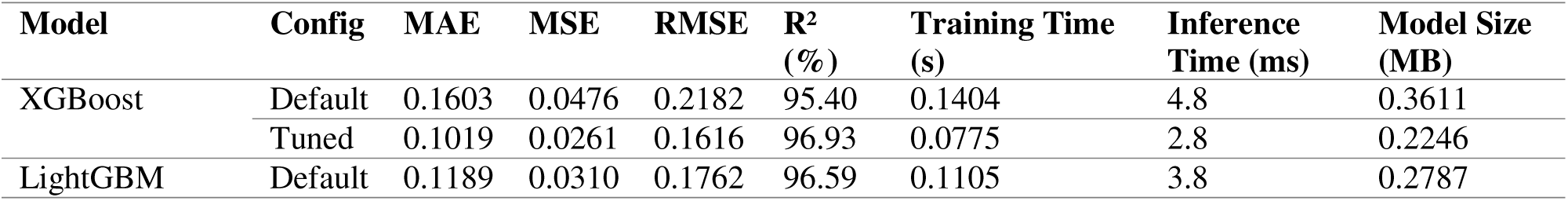
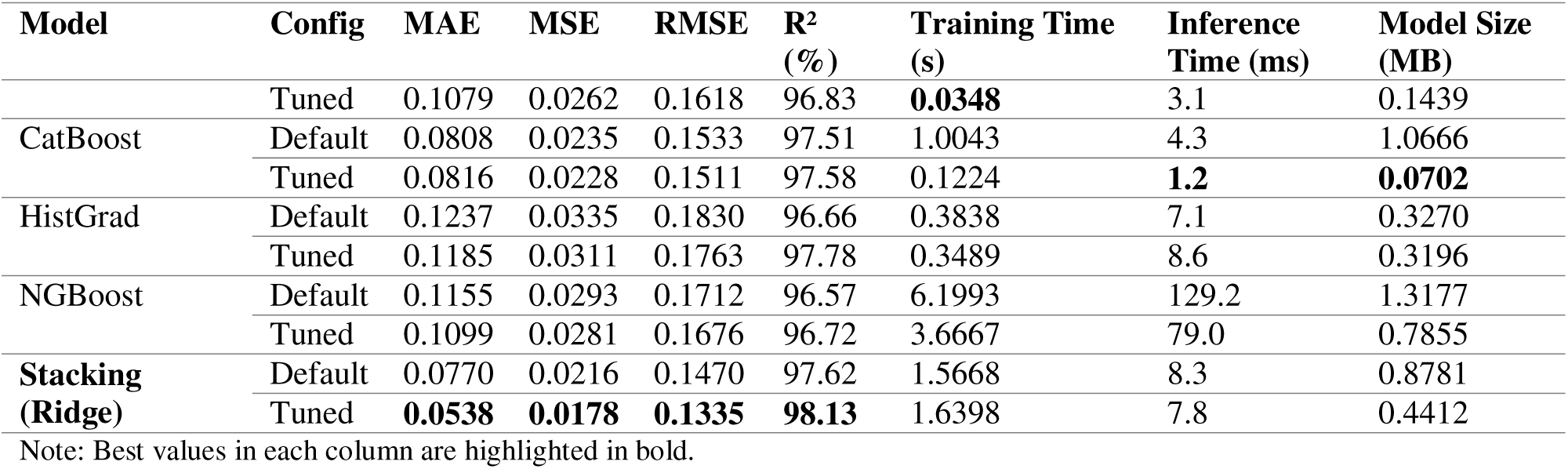
Performance comparison after FS.

### 4.4 Impact of FS on Model Performance

To assess the impact of FS on model performance, tuned model configurations were compared before and after FS. In particular, these models were selected because they provided more competitive baselines than default settings. Particularly, Table 7 reports four key metrics: CV R^2^ (predictive accuracy), training time (computational cost), inference time (deployment speed), and model size (memory footprint). Together, these metrics provide a balanced view of predictive and practical performance. In addition, Fig. 6 showed CV R^2^ measures before and after FS. Most models provide modest gains; notably, HistGrad improved the most, while the Stacking Ensemble consistently achieved the highest accuracy. Therefore, HistGrad gained 1.13 percentage improving from 96.65% to 97.78%, however, it required longer inference time and a larger model size. XGBoost improved slightly (0.11%) while reducing training time by 64%, inference time by 48%, and model size. The Stacking Ensemble remained the top performer, improving marginally from 98.12% to 98.13%, confirming its strong baseline. However, CatBoost maintained deployment efficiency, thereby reducing inference time by 40% (2.0 ms to 1.2 ms). As well, LightGBM showed consistent efficiency gains, with training time reduced by 33%, inference by 21%, and model size by 2%. Furthermore, training time was reduced the most for XGBoost, CatBoost, and Stacking Ensemble models. Meanwhile, inference speed improved notably for XGBoost and CatBoost. Model size remained stable or improved, except for HistGrad. It is worth noting that FS improved efficiency for most models while preserving competitive accuracy.

**Fig. 6.**
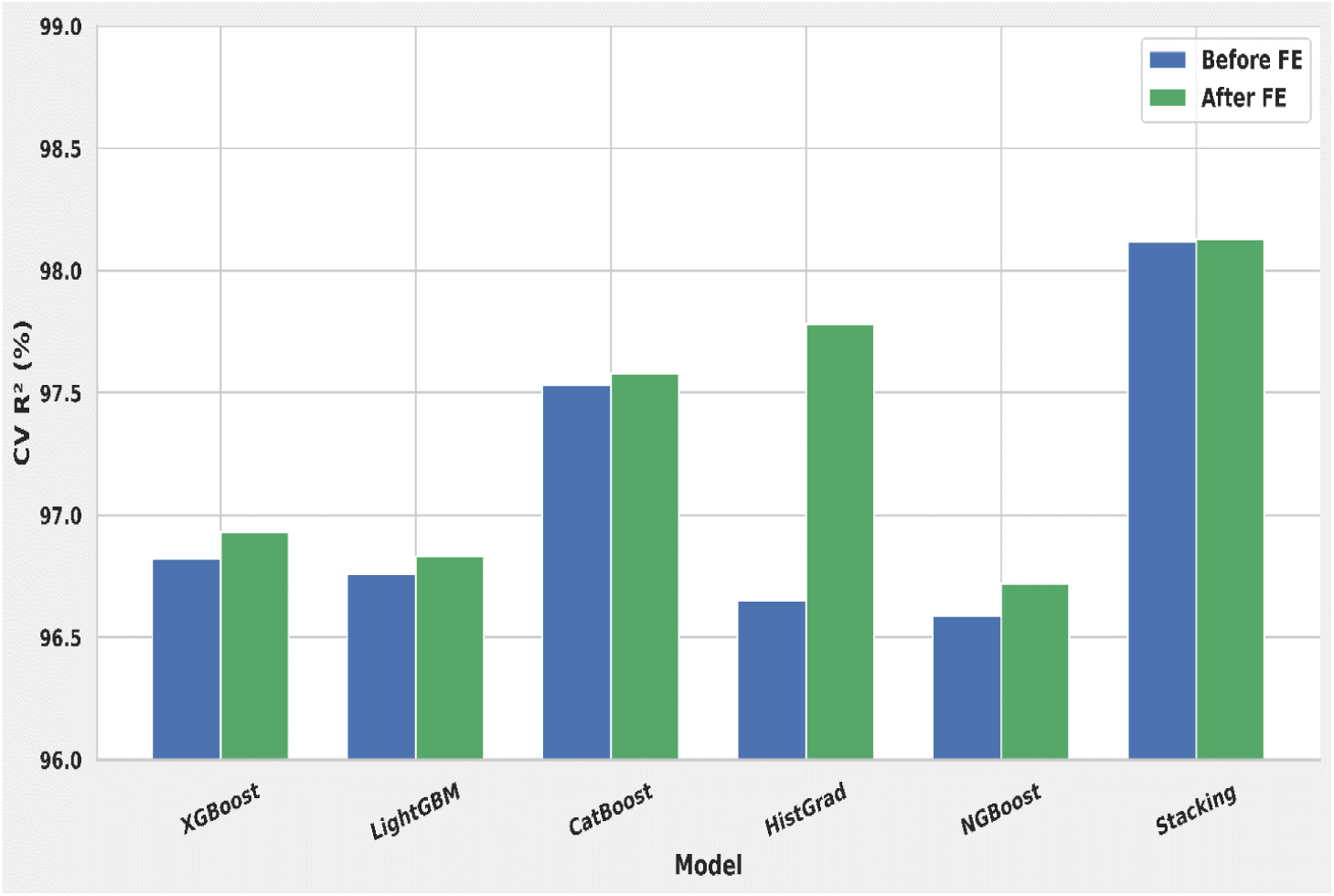
Effect of FS on Tuned Model Performance

**Table 7.** Impact of FS on tuned model performance.

| Tuned Model | Condition | CV R<sup>2</sup> (%) | Training Time (s) | Inference Time (ms) | Model Size (MB) |
| --- | --- | --- | --- | --- | --- |
| XGBoost | Before FS | 96.82 | 0.2175 | 5.4 | 0.2251 |
|  | After FS | **96.93** | **0.0775** | **2.8** | **0.2246** |
| LightGBM | Before FS | 96.76 | 0.0518 | 3.9 | 0.1466 |
|  | After FS | **96.83** | **0.0348** | **3.1** | **0.1439** |
| CatBoost | Before FS | 97.53 | 0.2371 | 2.0 | 0.0705 |
|  | After FS | **97.58** | **0.1224** | **1.2** | **0.0702** |
| HistGrad | Before FS | 96.65 | 0.2299 | 5.2 | 0.1906 |
|  | After FS | **97.78** | 0.3489 | 8.6 | 0.3196 |
| NGBoost | Before FS | 96.59 | 4.0905 | 83.4 | 0.7830 |
|  | After FS | **96.72** | **3.6667** | **79.0** | 0.7855 |
| **Stacking (Ridge)** | Before FS | 98.12 | 3.5934 | 12.9 | 0.4449 |
|  | After FS | **98.13** | **1.6398** | **7.8** | **0.4412** |

Note: Best values in each column are highlighted in bold.

Finally, the tuned Stacking Ensemble was chosen as the final model for CVD risk prediction. It achieved the highest CV R^2^ of 98.13% with the lowest error metrics and notable efficiency gains.

### 4.5 Model interpretation with SHAP

To improve interpretability, SHAP analysis was applied to the post FE-tuned Stacking Ensemble regressor. Fig. 7 presents the SHAP global summary plots of this final regressor. Therefore, X-axis shows SHAP values (feature impact on predictions), while Y-axis ranks feature by importance. Moreover, the color gradient from red (high values) to blue (low values) showed how feature importance influences the direction of model prediction. Indeed, SHAP analysis indicated interpretable feature importance using the eleven selected predictors. BMI, TC, and DMST remained the top contributors, along with consistent SHAP values around –0.90. In addition, SBP, LDL, HDL, PA, DBP, BPCAT, WHtR, and SMK showed uniform impacts of approximately ±0.90. From the analysis, it is shown that high values of BMI, cholesterol fractions, and blood pressure contributed as increasing factors in CVD prediction measures. Conversely, HDL and PA exerted protective effects. The removal of low-importance variables (SEX, FHxCVD, FBS, AGE, AC) simplified the model and improved interpretability. Overall, the analysis confirms that hypertension, cholesterol, age, and BMI are the primary drivers of CVD risk prediction.

**Fig. 7.**
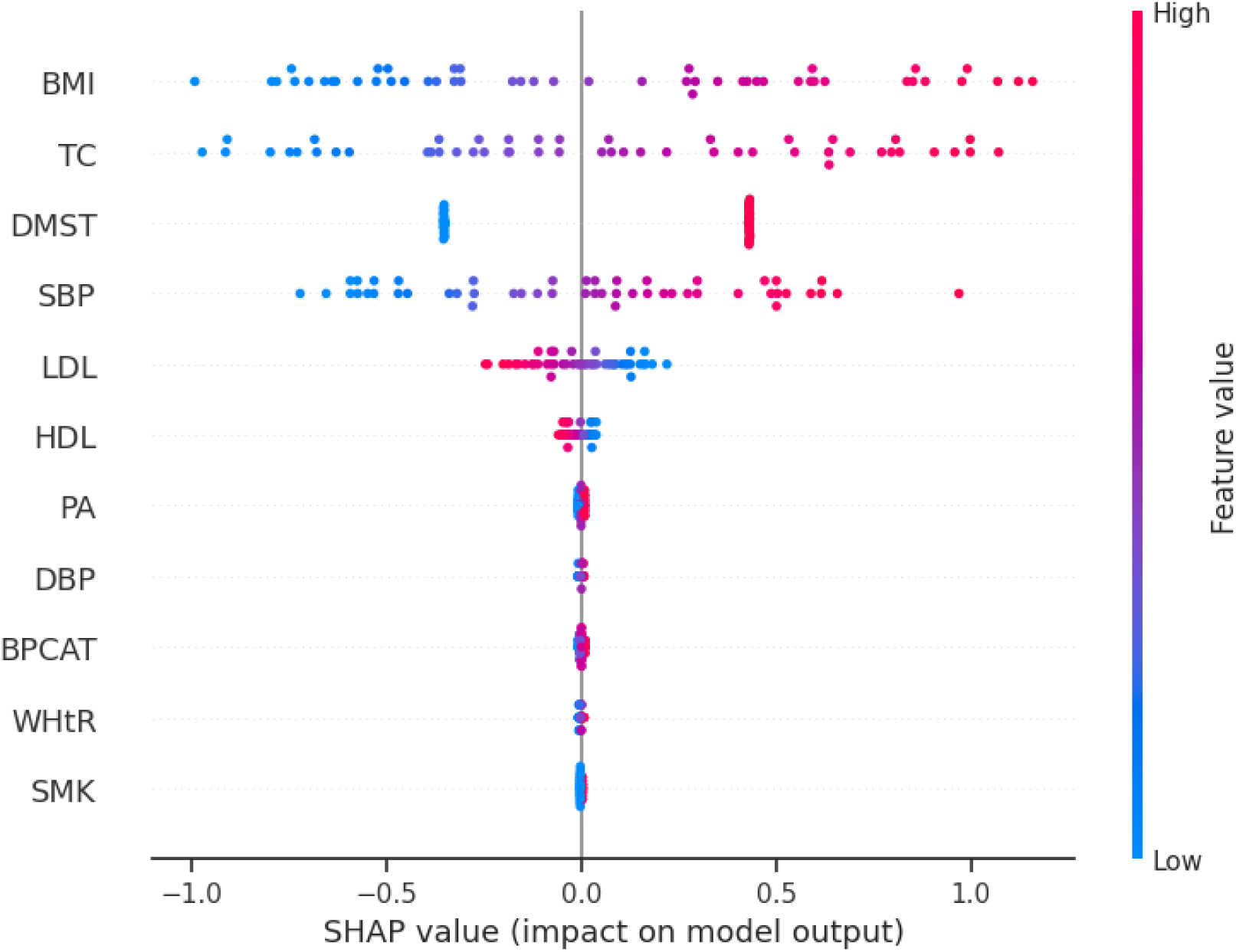
SHAP summary plots

Thus, SHAP analysis showed that FS conserved clinically meaningful predictors. It aligned model outputs with known cardiovascular risk factors. As a result, it improved transparency for clinical use.

### 4.6 Comparison with Existing Studies

A comparative analysis between prior research and our proposed approach for CVD regression is presented in Table 8. However, most reviewed studies did not incorporate XAI, efficiency metrics, CV adoption, consensus-based FS or practical deployment through web applications. In addition, our study applied a 10-fold CV to ensure robust and reliable outcomes. It achieved good performance with 98.13% R^2^. In addition, our approach integrated SHAP based explainability, used consensus feature selection across five methods, evaluated computational efficiency, and showed deployment through web platforms. Overall, these contributions highlight the distinctiveness and clinical relevance of our CVD framework in real-world settings.

**Table 8.** Comparison summary of our study with recent literature.

| Study | Model | Dataset Size | Performance | CV Strategy | XAI | FE | Efficiency Metrics | Practical Deployment |
| --- | --- | --- | --- | --- | --- | --- | --- | --- |
| [17] | Random Forest | 112 | 95% ($R^2$) | No | No | No | No | No |
| [37] | Gradient Boosting | 70,000 | 74% (Acc) | Yes | No | Yes | No | No |
| [38] | Random Forest | 518 | 85.01% (Acc) | No | No | No | No | No |
| [39] | Stacking | 8734 | 87.8% (Acc) | Yes | No | Yes | No | No |
| [40] | Random Forest | 907 | 71.1%(AUC) | Yes | Yes | Yes | No | No |
| [41] | Voting | 70,303 | 73.87%(Acc) | Yes | No | No | No | No |
| [42] | Gradient Boosting | 8,495 | 76.2% (Acc) | No | No | Yes | No | No |
| [18] | Stacking | 1,529 | 96% ($R^2$) | Yes | Yes | No | No | No |
| [43] | Stacking | 2,066 | 0.0904 (MSE) | Yes | No | No | No | No |
| Present study | Stacking Ensemble | 1,529 | 98.13% ($R^2$) | Yes | Yes | Yes | Yes | Yes |

### 4.7 Web Visualization

The web application provides role-specific dashboards tailored to different user types. In particular, Lab assistants can manage clinical tests through the test management interface shown in Fig. 8. They can add or update patient test results, including glucose levels, blood pressure, and haemoglobin counts. Moreover, the interface allows them to maintain the test catalogue provided by their institute by adding new tests or removing outdated ones. On successful authentication, patients are redirected to the personalized patient dashboard. Therefore, they can access their medical records and history. Meanwhile, doctors are directed to the dedicated doctor dashboard, which provides AI powered diagnostic tools and patient management options.

**Fig. 8.**
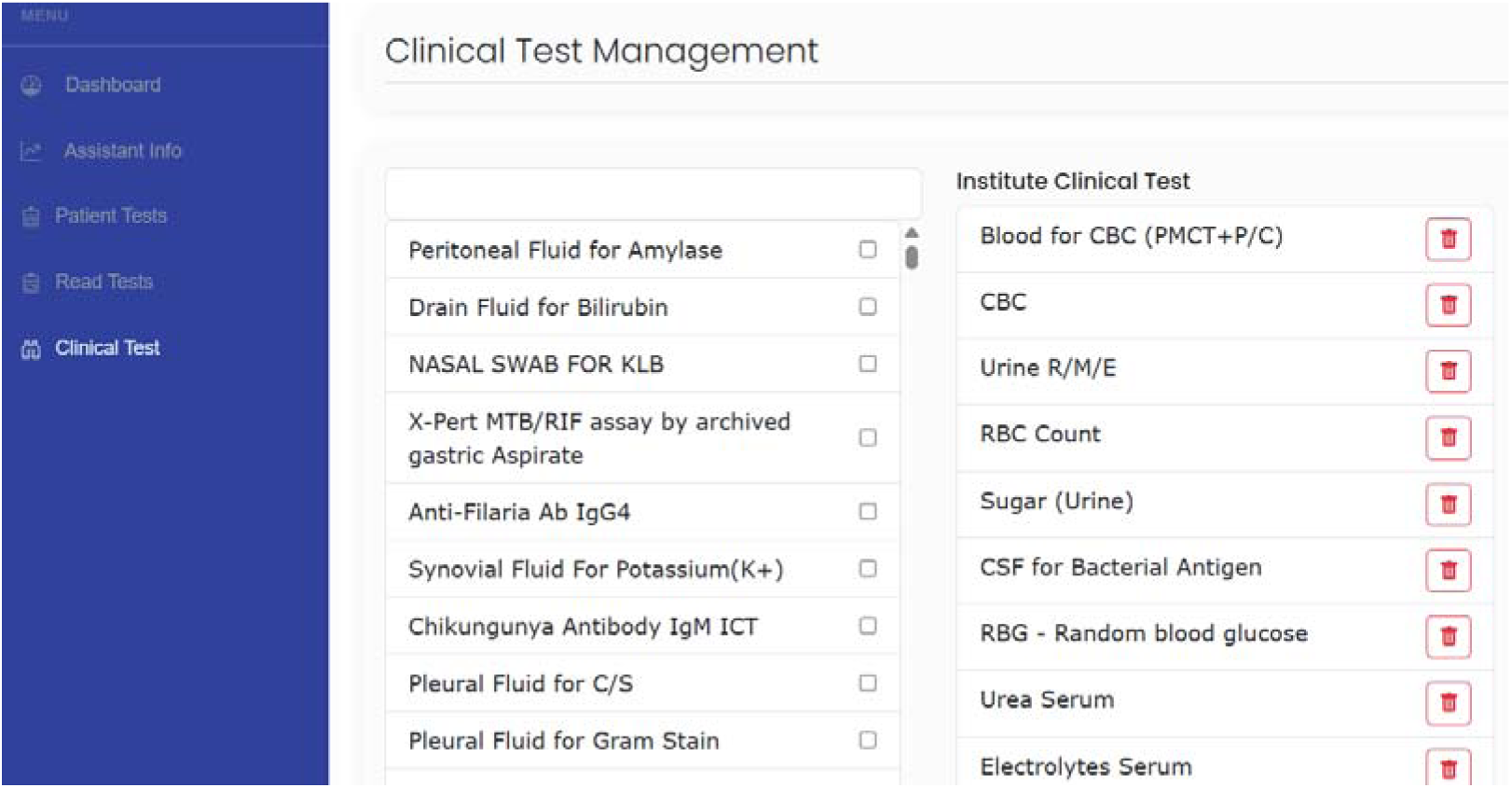
Clinical test management

The web tool interface displays input fields for demographic and clinical parameters such as age, gender, blood pressure, cholesterol levels, and other cardiovascular indicators. Based on these inputs, the web application generates predictions, as shown in Fig. 9. It also provides CVD risk score classification ranging from very low to very high in accordance with WHO standards [44].

**Fig. 9.**
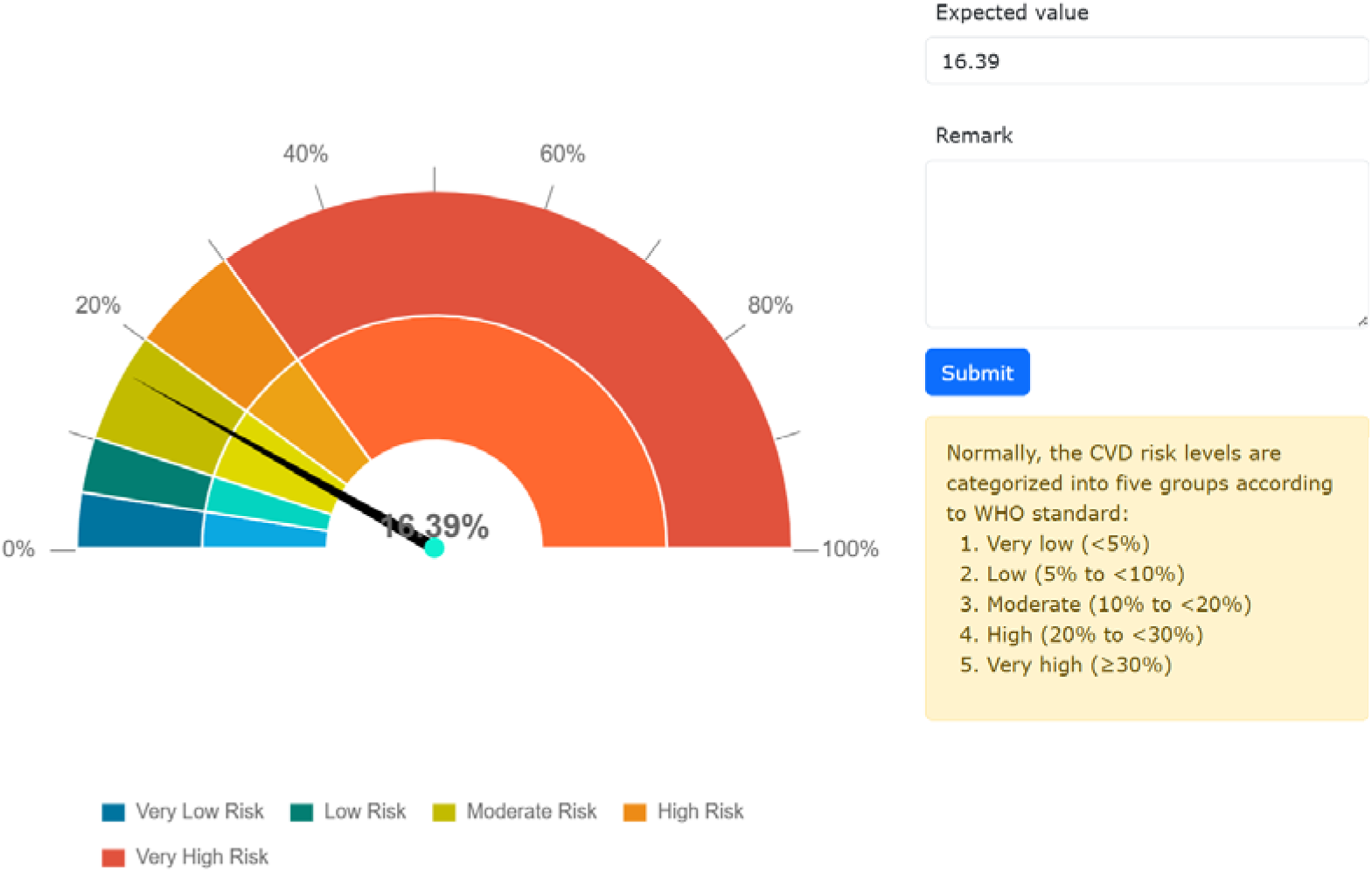
Visualization of CVD risk prediction

Overall, the visualization of the web application prioritized simplicity and usability. It used straightforward language and smooth navigation, which ensured accessibility for individuals with limited digital literacy. Importantly, the tool has been tested and validated in real-world clinical settings at Asgar Ali Hospital, Dhaka, Bangladesh (approval letter in supplementary sections) to demonstrate its practical applicability and reliability.

## 5 Conclusions and Future Work

This paper introduced an ML-powered CVD management system using a stacking ensemble regressor that integrates adaptive inference-based gradient-boosting models. The proposed framework showed robust, reliable performance in predicting CVD risk scores. Moreover, an XAI technique, SHAP, was integrated to provide transparency in the model and insights into the most important features that lead to clinical decisions. This CVD risk score framework was applied to successfully handle missing data effectively by the use of MICE, and then the relevant features were selected with consensus-based FS techniques. Next, the system was deployed for practical use with the model applied via an ML Flask API and a Laravel-based web platform. Experimental results showed that for a dataset of 1,529 patient records with 22 features, after feature selection, the tuned Stacking Ensemble achieved the best overall performance with a CV R^2^ of 98.13% and the lowest error metrics. In addition, it was found that CatBoost had the best deployment efficiency, offering fast inference and small model size, while LightGBM had the lowest training-time with competitive accuracy.

In the future, the research can be extended to incorporate longitudinal patient data and real-time monitoring that will strengthen clinical decision-making. Besides, consideration of deep learning models and medical image processing techniques can further improve early detection capabilities.

## Data Availability Statement

The dataset used in this study is publicly available at the following repository: https://data.mendeley.com/datasets/d9scg7j8fp/1

