## Supplemental Files for "A Consensus-Driven Stacking Ensemble Framework for Interpretable Cardiovascular Risk Prediction and Clinical Deployment"

**Table of Contents**

S1. Exploratory Data Analysis

S1.1 Feature Distributions

S1.2 Pearson Correlation Heatmap

**S1. Exploratory Data Analysis**

This section presents the exploratory data analysis (EDA) conducted on the dataset used in this study (n = 1,529 patient records). Supplementary Figs. S2a–g show the univariate distributions of continuous clinical features, while Supplementary Figs. S2h–l show the distributions of categorical variables. Supplementary Fig. S3 presents the Pearson correlation heatmap illustrating linear associations among all features and the target variable (CVDRISK). These analyses informed preprocessing decisions including outlier removal via IQR, MICE imputation, and consensus-based feature selection.

**S1.1 Feature Distributions**

The distributions of continuous variables are presented in Figs. S2a–g and categorical variables in Figs. S2h–l. BMI is moderately right-skewed, with most participants falling in the normal-to-overweight range. DBP follows a near-symmetric distribution, while SBP shows greater dispersion, reflecting population-level hypertension variability. TC and FBS display mild right skewness, indicating a subset with elevated cardiometabolic risk markers. HDL is approximately balanced, whereas LDL exhibits wider spread, consistent with inter-individual lipid variability. Categorical variables (Figs. S2h–l) are roughly balanced across sex, smoking status, and family history of CVD, supporting unbiased group comparisons.

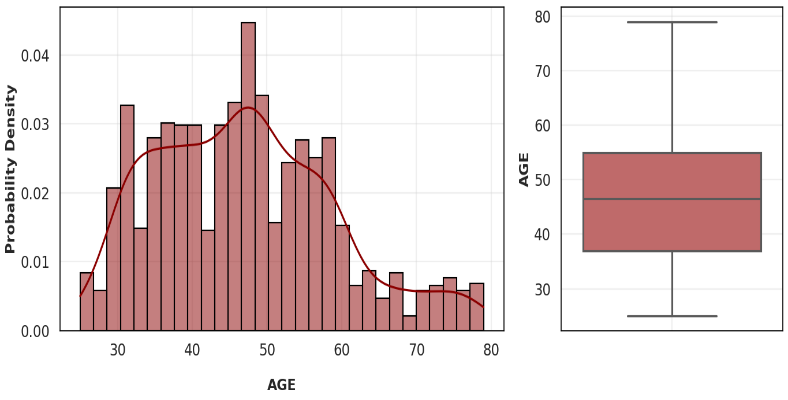

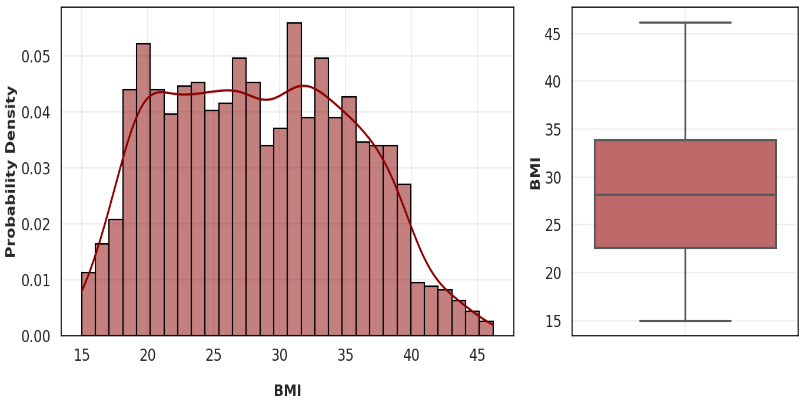

(a)	(b)

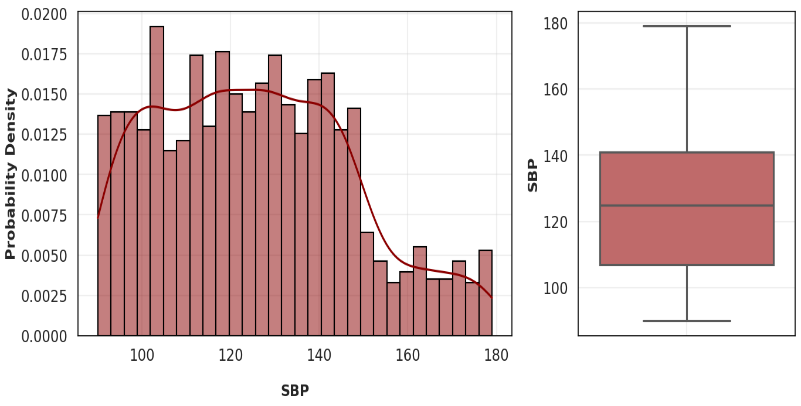

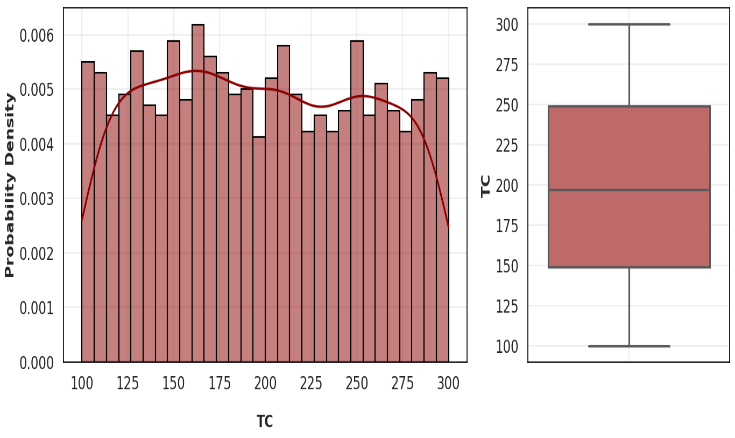

(c)	(d)

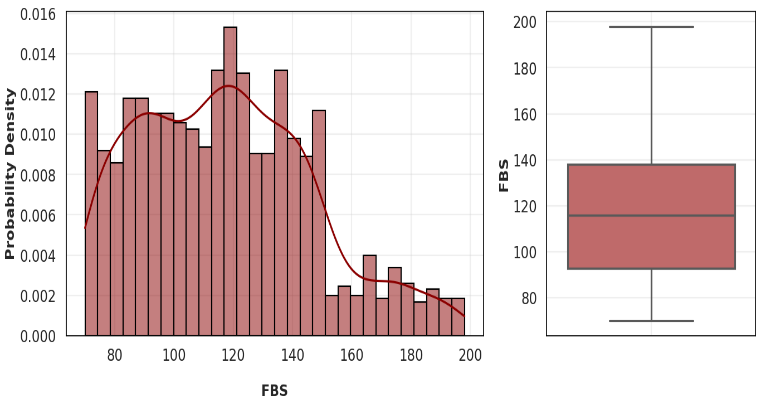

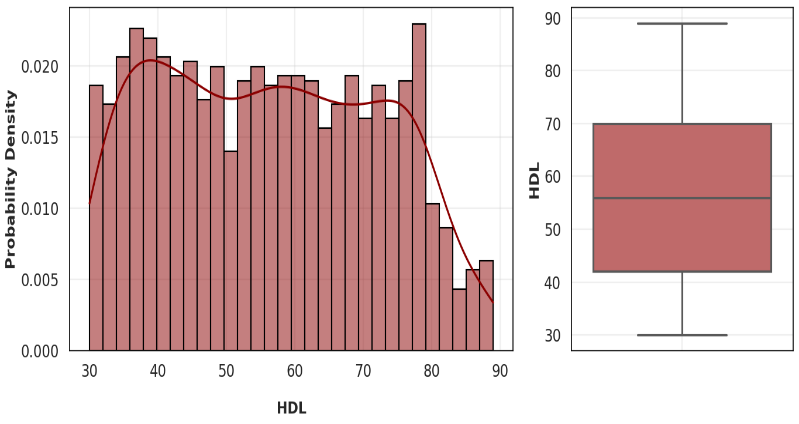

(e)	(f)
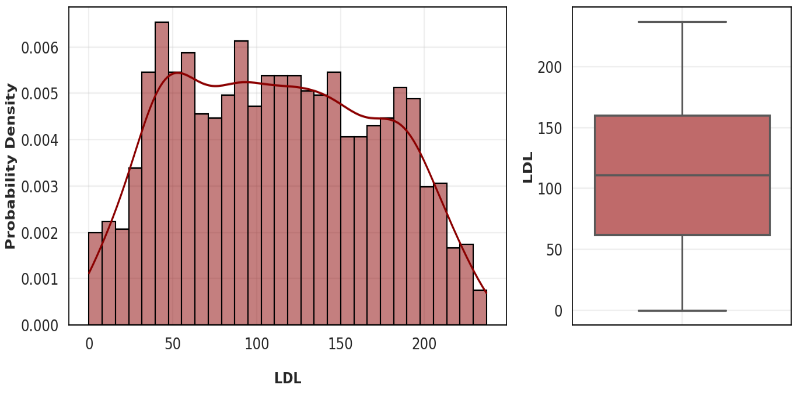

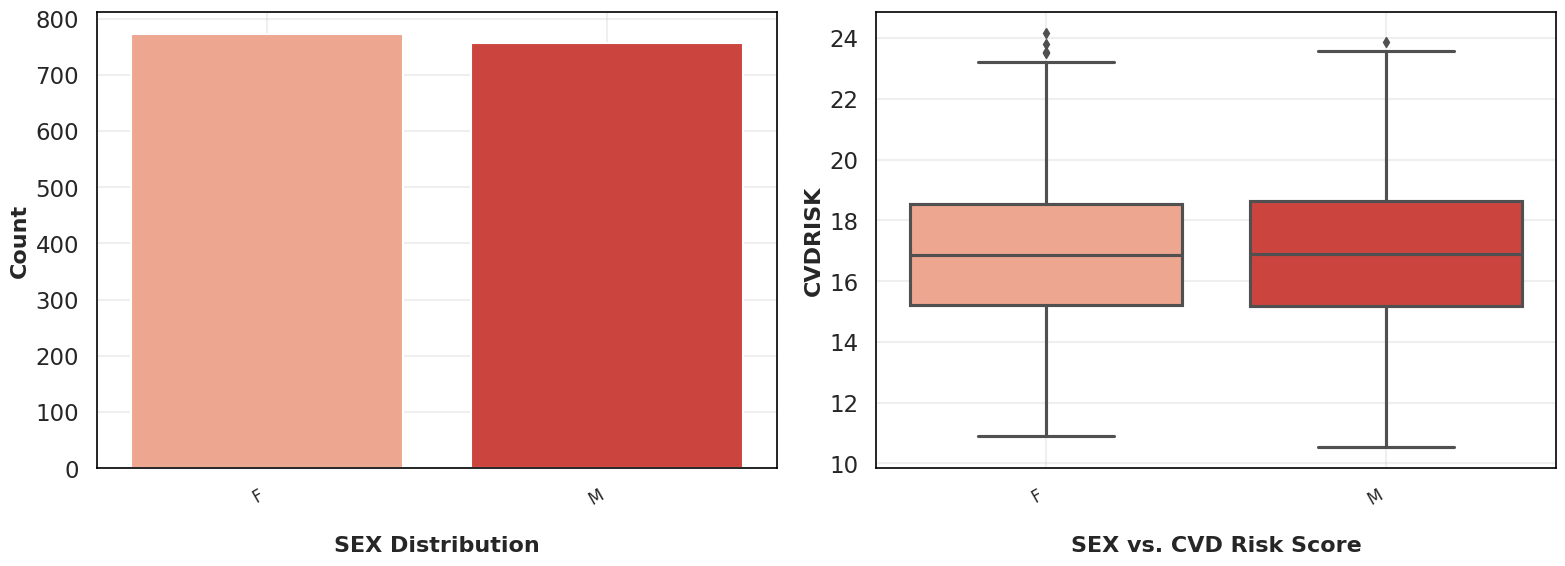

(g)	(h)

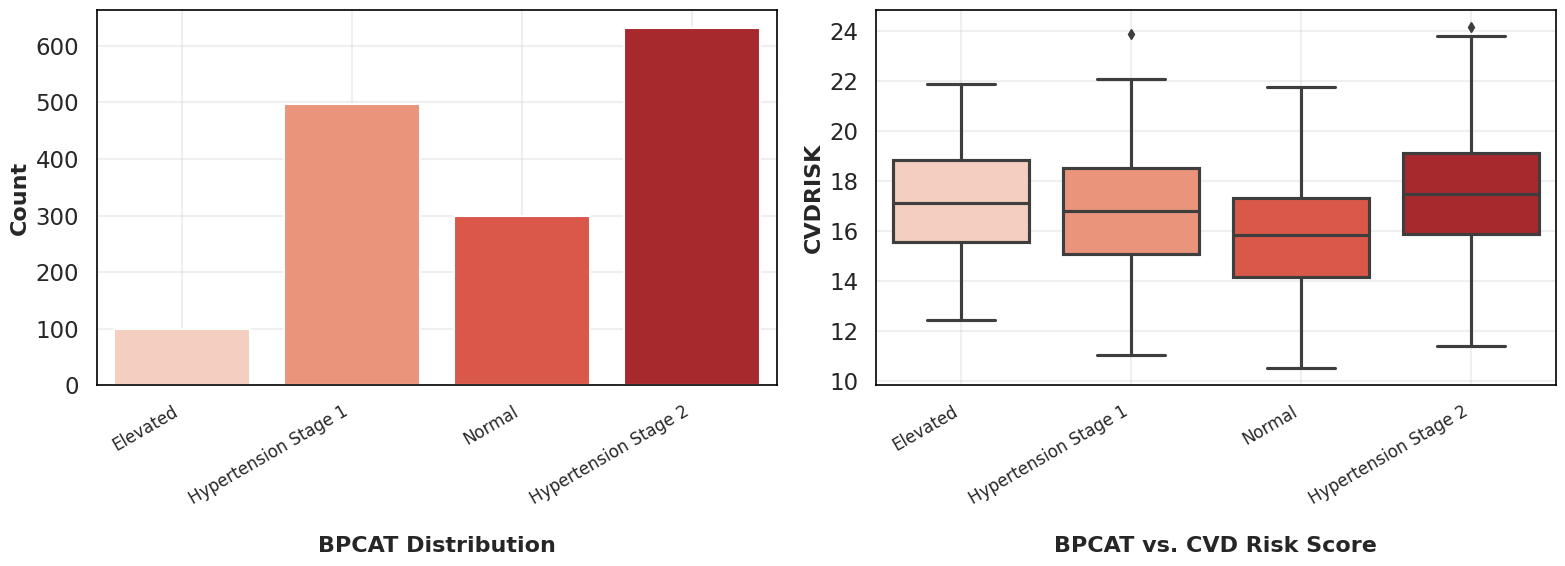

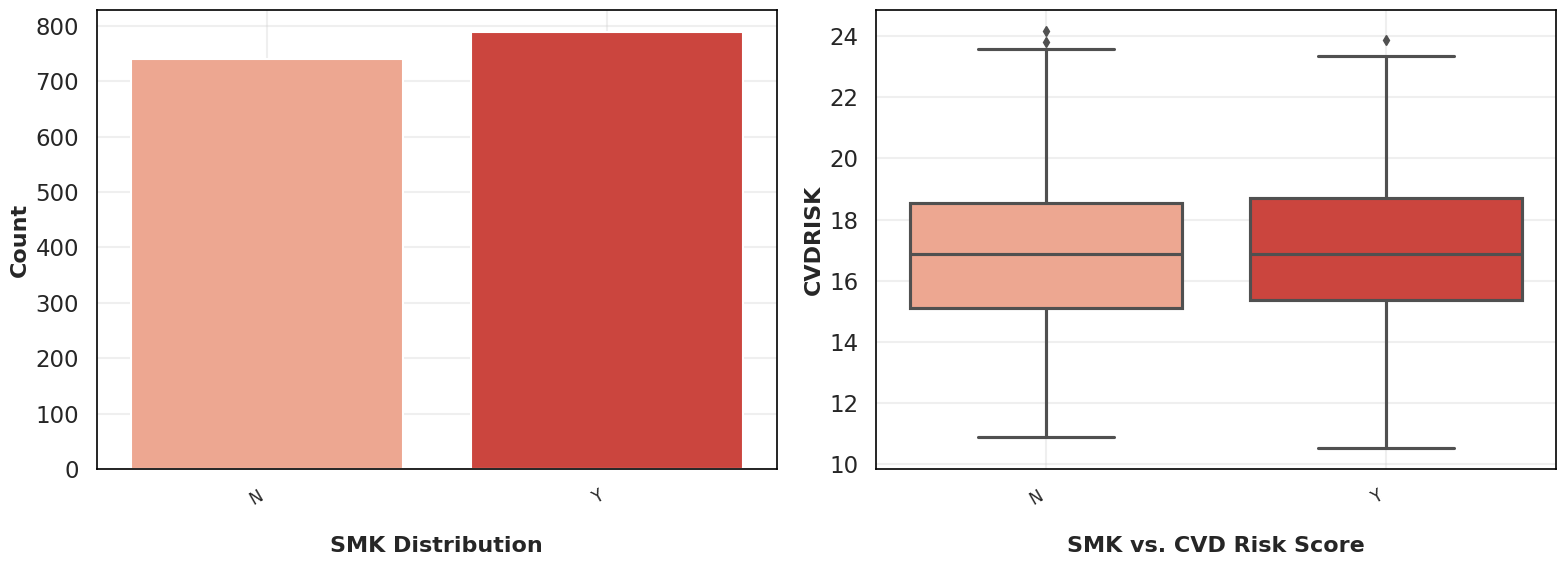

(i)	(j)

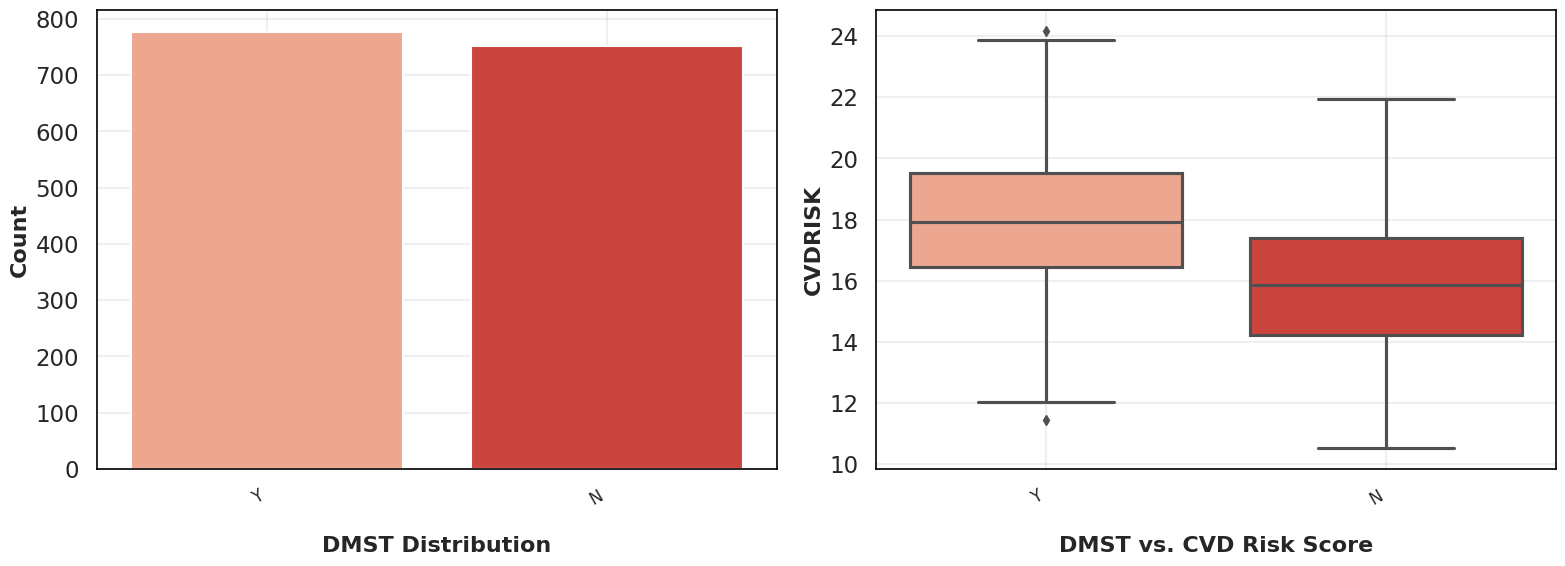

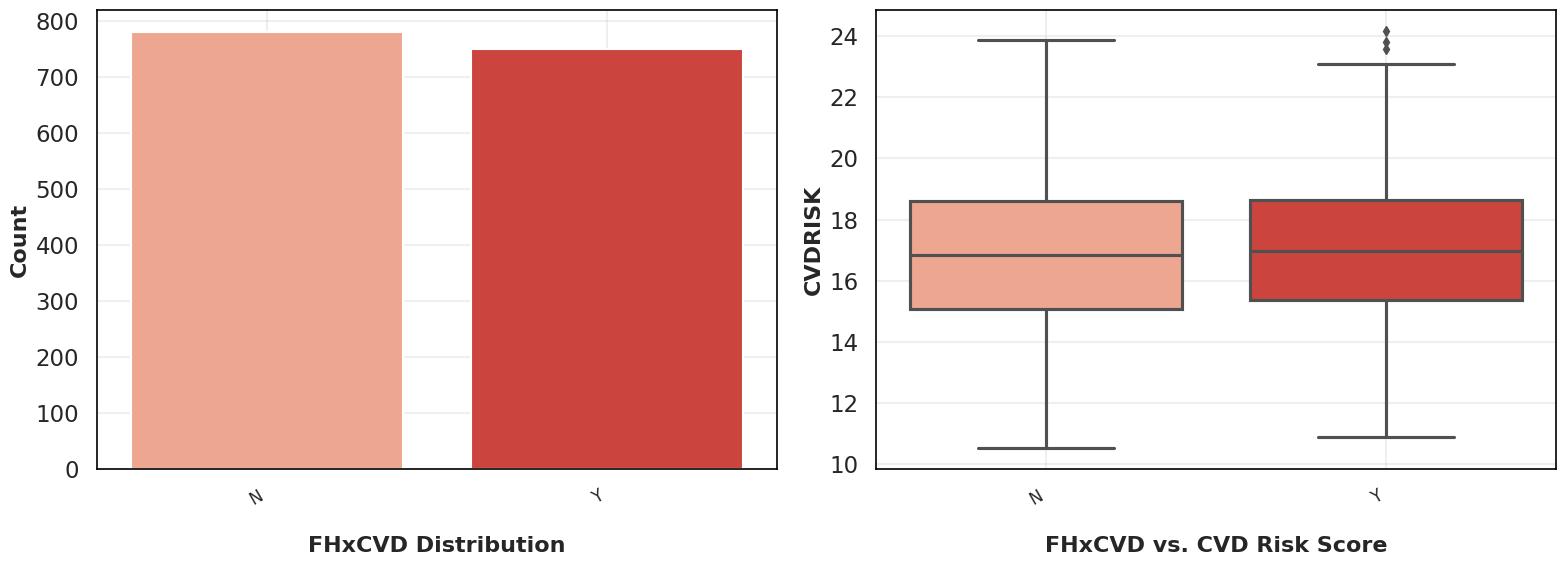

(k)	(l)

*Fig. S2. Feature distributions of the CVD risk score dataset (n = 1,529). Continuous variable histograms: (a) Age (AGE), (b) Body Mass Index (BMI), (c) Abdominal Circumference (AC), (d) Waist-to-Height Ratio (WHtR), (e) Systolic Blood Pressure (SBP), (f) Diastolic Blood Pressure (DBP), (g) CVD Risk Score (CVDRISK). Categorical variable bar plots: (h) Sex (SEX), (i) Blood Pressure Category (BPCAT), (j) Smoking Status (SMK), (k) Physical Activity Level (PA), (l) Family History of CVD (FHxCVD). Categorical variables are coded as: 1 = presence/yes, 0 = absence/no.*

**S1.2 Pearson Correlation Heatmap**

Figure S3 presents the Pearson correlation heatmap for all numerical features relative to the CVD risk score target variable (CVDRISK). BMI demonstrates the strongest positive association (r = 0.62), followed by SBP (r = 0.48), confirming their role as primary predictors. BPCAT and TC show moderate positive correlations, while HDL exhibits a weak negative association, consistent with its established cardioprotective role. FBS, LDL, and DBP show moderate correlations, supporting their retention in the feature pool. These findings directly validated the consensus-based feature selection outcomes reported in Table 3 of the main manuscript.

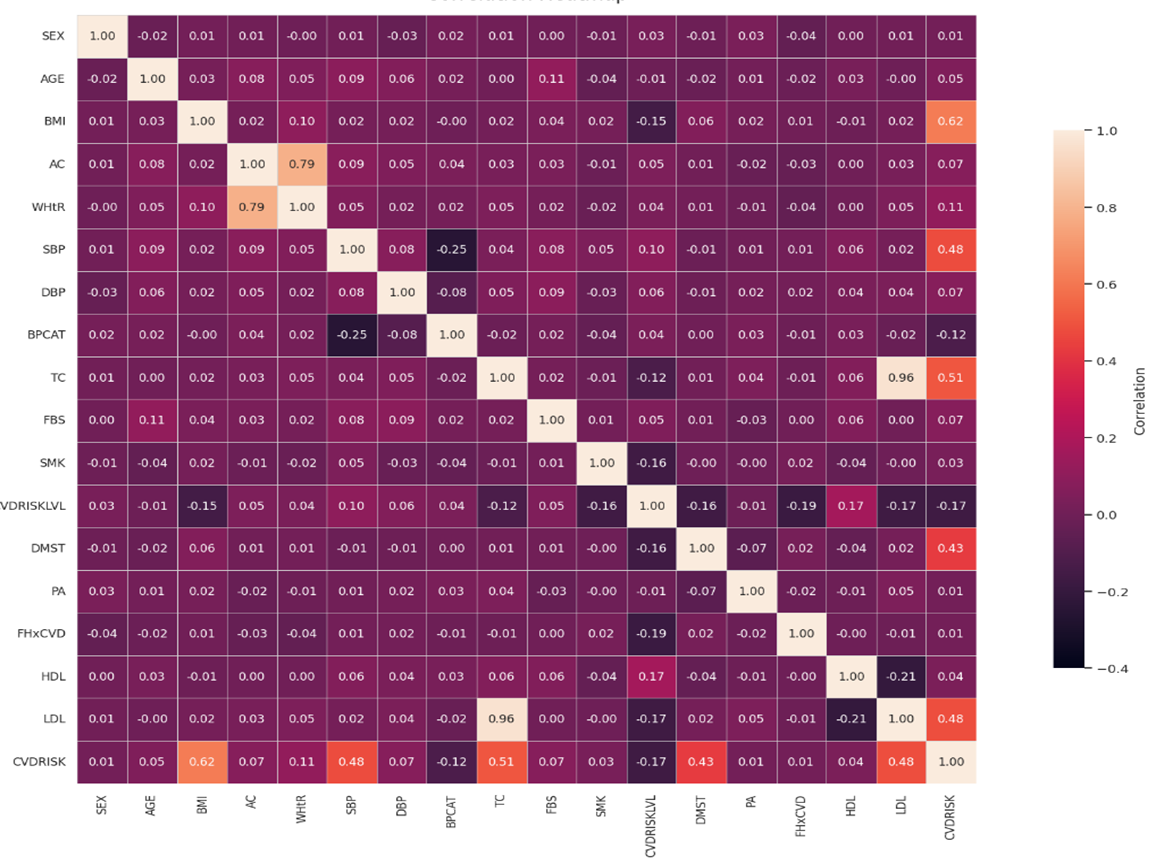

***Fig. S3.*** *Pearson correlation heatmap of the CVD risk score dataset. Cell values represent pairwise Pearson correlation coefficients (r) between all numerical features. The colour scale ranges from strong negative (blue) to strong positive (red) correlations. Features with |r| ≥ 0.40 with CVDRISK are considered strong predictors and were prioritized in the consensus-based feature selection process (Section 3.4).*
